# Wildfire pollution exposure during childhood adversely affects cognitive and neural development

**DOI:** 10.64898/2026.06.08.26355154

**Authors:** Nicholas Judd, Rogier Kievit

## Abstract

Air pollution has well-documented negative cardiovascular and respiratory consequences. However, the impact of fine particulate matter pollution (PM) on brain development is unclear. Animal studies suggest that exposure to early-life PMcan cause adverse neurodevelopmental outcomes, but in vivo human work has been hampered by cross-sectional designs and heavily confounded PM_2.5_ exposure measures. Here we use an innovative natural experimental design to isolate the effects of wildfire pollution on neurocognitive development in a large cohort of children (N>9000, 4 waves, age 9-16). Doing so, we find that greater wildfire PM_2.5_ exposure is robustly associated with slower brain development and shallower cognitive improvement across early adolescence. Our study underscores the urgent public health concern that wildfire PM_2.5_ poses for childhood development.

## Introduction

Air pollution has serious health consequences. Small particulate matter in particular (e.g., < 2.5 micrometers; PM_2.5_) has been identified as especially harmful, estimated to cause the premature deaths of ∼4 million people per year (*1*, *2*). In addition to well-documented cardiovascular and respiratory impacts, a growing body of animal work has highlighted the effects of particulate matter pollution on the central nervous system (*3–5*). PM_2.5_ can enter the brain directly via the olfactory and trigeminal nerves, or indirectly, passing through the alveoli into the bloodstream (*1*, *4*). Particulate matter has been found to accumulate in human blood, brain, cerebral spinal fluid, and the fetal side of the placenta (*6–8*). After entry, animal and post-mortem human work indicate PM_2.5_ exposure causes oxidative stress, systemic neuroinflammation, and vascular damage, leading to neurodegeneration (*3*, *4*, *9*, *10*).

The adverse effects of air pollution are likely even more severe during neurodevelopment (*3*, *11*)–an extended period of dynamic neural changes at multiple levels (cellular, circuit, and system), accompanied by concurrent behavioral, psychiatric, and cognitive changes (*12–15*). This heightened level of neural plasticity in early life is hypothesized to increase the salience of environmental influences and contribute to the fetal programming of neurodevelopmental disorders (*5*, *11*, *14*, *16*, *17*). Children demonstrate marked interindividual differences in developmental trajectories (*18*, *19*), which persist and manifest in later-life adult outcomes (*20*, *21*). In addition to heightened vulnerability, children also experience greater exposure to outdoor pollution because they are more likely to be outdoors, engage in more exercise, and have higher respiratory rates (*22*, *23*). One of the only longitudinal studies to date (*24*) found neighborhood PM_2.5_ pollution to be associated with neurite development across the cortex of children (*23*). For these reasons, it is vital to understand the role early-life PM_2.5_ exposure plays in shaping neurodevelopment trajectories.

Despite plausible biological mechanisms and compelling evidence from experimental animal studies, the effect of PM_2.5_ pollution on human neurobiology is far less clear. This is in large part due to ethical constraints and the practical challenges of studying long-term outcomes in humans in vivo. For this reason, almost all prior human neural work on air pollution uses *neighborhood* PM_2.5_ pollution (*24*). Unfortunately, these measures are heavily confounded with a myriad of other socioeconomic and individual-level characteristics known to be associated with neural and cognitive outcomes (*16*, *25*, *26*). This tight coupling is driven by both implicit economic mechanisms (e.g., house prices) and explicit prior policy choices (e.g., differential permitting of highways/industry), overwhelmingly affecting less affluent, usually minority, communities (*16*, *27*). Previous work (*25*, *27*) has established that families with lower incomes tend to live in neighborhoods with higher air pollution. This stratification is best exemplified by the well-established genetic correlations between one’s neighborhood, cognitive performance, and neural development (*28–31*). In addition to individual-level sorting, neighborhood-level characteristics are not equal– neighborhoods with worse air pollution also tend to be chronically underfunded with fewer good schools, worse healthcare access, limited access to services, and higher crime rates (*27*, *32*), all of which are also associated with worse life outcomes. This constellation of (dis)advantage makes it extremely difficult to isolate the impact of PM_2.5_ pollution in humans. Here, we leverage a unique source of particulate matter– wildfire-specific PM_2.5_ pollution (*33*) – allowing us to uncouple it from these various confounders.

Wildfires can be seen as an example of a natural experiment– a ‘random-like’ exogenous external event less affected by confounds usually associated with observational analysis. Unlike neighborhood pollution, which is often caused by local settings (e.g., road traffic, industry), wildfire pollution affects different neighborhoods equally and is uncorrelated with SES in the United States (*33*, *34*). This allows us to isolate, in principle, a more causal estimate of air pollution exposure in humans. Natural experimental designs have been applied to epidemiological and behavioral outcomes with great success– establishing causal links between air pollution exposure and childhood respiratory hospitalizations or mortality (*1*, *35*). Despite their potential to reveal the effects of early-life exposure on subsequent development in humans, natural experimental designs have been largely neglected in neuroscience (although see (*36*, *37*)). Previous work in non-human primates compared infant macaques born directly before and after severe wildfire pollution, finding that exposed infants showed greater inflammation, blunted cortisol, more passive behavior, and memory impairment (*38*). This finding of memory impairment is consistent with a cross-sectional study in children, which demonstrated that cumulative childhood wildfire pollution the year before decreased standardized test scores (*39*).

Wildfires are a useful design not just because they can provide stronger causal and mechanistic evidence of pollution’s adverse effects on neurobiology, but also for two other reasons. First, mounting evidence suggests human-induced climate change has increased wildfire size, severity, and prevalence (*40*)– to such an extent that increased wildfire pollution threatens to reverse positive trends of ambient air quality in the United States (*41*). Second, the chemical composition of wildfire PM_2.5_ pollution is also of concern, due to higher concentrations of toxicants, microbes, carcinogenic hydrocarbons, and heavy metals (*42–44*). Together, the urgency to understand the impact of wildfires on childhood development is clear: the increased prevalence and toxicity of wildfire pollution, paired with children’s heightened neural vulnerability and increased exposure to air pollution, is likely to have a profound impact on their development. To the best of our knowledge, no studies have examined how (or if) wildfire pollution adversely affects human brain development.

Many major life factors – such as exposure to pollution – are either infeasible or unethical to study experimentally in humans. Fortunately, recent advances in methodology and design allow causal inference in otherwise observational phenomena under certain assumptions (*45*). Here, we focus on wildfire-specific pollution and within-person change over adolescence, to isolate how prior PM_2.5_ pollution shapes human neural development. Our innovative approach was achieved by merging two rich open-access resources with overlapping geographic and temporal characteristics: *1)* daily 10km grid estimates of wildfire-specific PM_2.5_ pollution in the contiguous United States (*46*), with *2)* a large longitudinal (4 waves over 22 sites) neuroimaging cohort (*47*). We develop perinatal and childhood-specific wildfire exposure measures to assess how prior exposure affects cognitive and neural developmental trajectories during adolescence. Based on experimental findings, we hypothesize that prior wildfire pollution will have wide-ranging, negative developmental effects on cognitive performance and structural neuroimaging outcomes.

## Results

To examine the impact of wildfire pollution on neurodevelopment, we match children from the largest longitudinal neuroimaging cohort in the world (ABCD v6; starting N = 9806) (*47*) to daily local-level PM_2.5_ wildfire smoke estimates for the contiguous United States(*46*). We develop two non-overlapping wildfire PM_2.5_ exposure measures per child (Figure 1a & 1b): one for the perinatal period (PSE: conception through 3 months of age) and another for cumulative exposure over the rest of childhood (CSE: 3 months through the first assessment date; ∼10 years old). Since the perinatal period is shorter than childhood, these two wildfire PM_2.5_ measures differ in their average exposure levels (140 vs 1145 PM_2.5_ μg/m^3^, respectively). We separate perinatal exposure for two reasons: perinatal development is a time of remarkable neural plasticity (*13*, *14*), leading to strong theoretical claims (*17*, *48*) and evidence (*49*, *50*) that early environmental exposures have a lasting effect on development. Our design takes advantage of the large temporal variation around the first assessment date (range 26-months) and the birthdate recruitment eligibility window (range 49-months)– allowing both wildfire-PMexposure measures to demonstrate large within-site variability and a minimal correlation with each other (Pearson’s r = 0.07; Sup. Figure 1 & 2).

**Figure 1:**
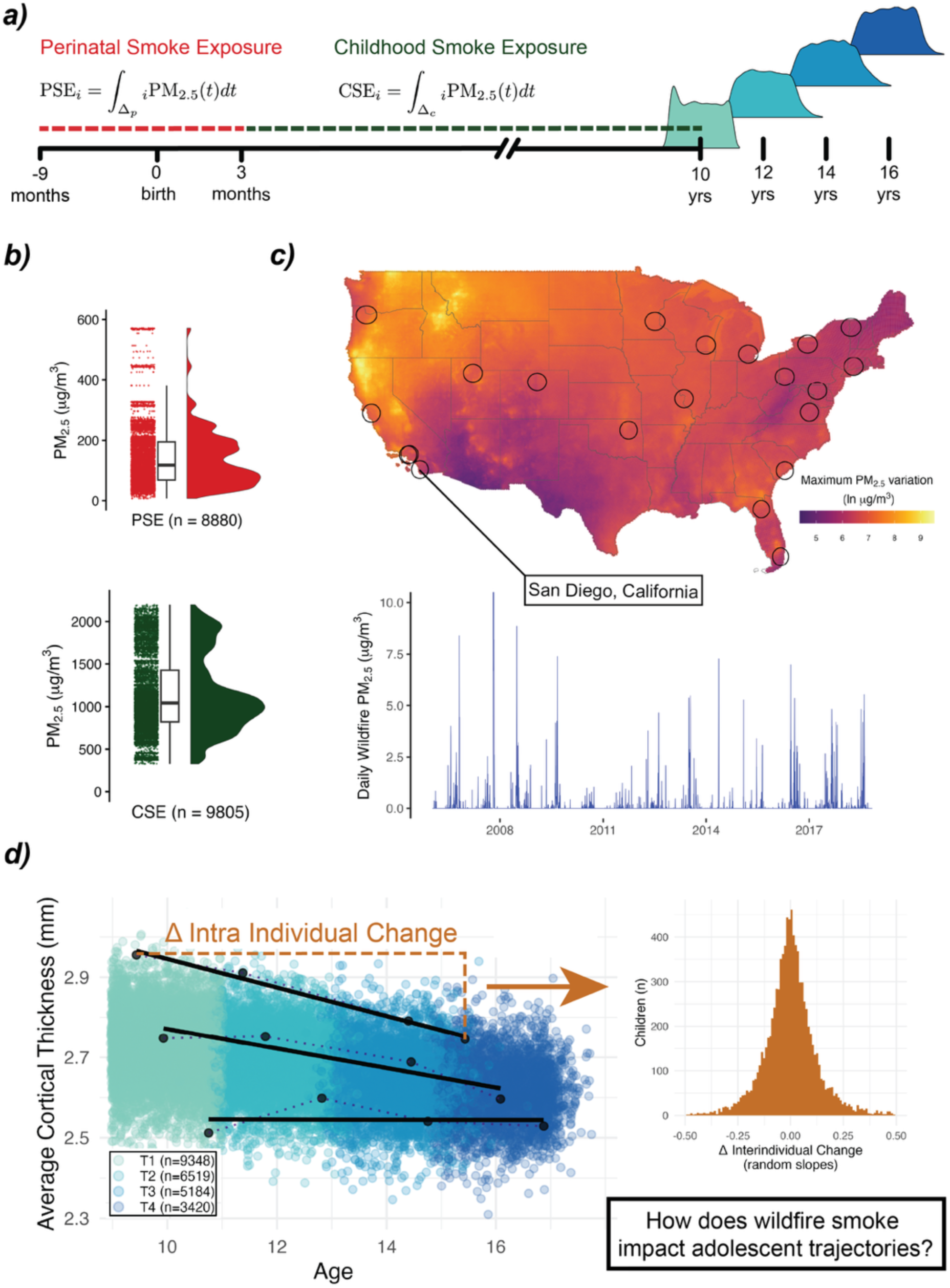
An overview of our study design in which we combined high-resolution daily wildfire-PM2.5 smoke pollution data with a large longitudinal neuroimaging cohort (ABCD; n = 9805), allowing us to isolate the impact of wildfire pollution on different adolescent trajectories (intra-individual change). First, **a)** we construct two smoke exposure measures per child, **b)** using high-resolution smoke pollution data. Perinatal Smoke Exposure (**PSE**) reflects a child’s cumulative exposure from conception up until 3 months of life; Childhood Smoke Exposure (**CSE**) was defined as a subject’s cumulative exposure from 3 months of age up until the first assessment date (i.e., Visit #1; ∼10 years of age). In **c)**, to illustrate high-resolution smoked pollution data we calculated the maximum PM_2.5_ 2.5 variation possible in 10km geospatial grids in the contiguous United States, defined as the difference in exposure between the earliest possible date of birth with the latest possible visit date (maximal exposure) and a subject with the latest possible date of birth and the earliest visit date (minimal exposure) and plot them in log units. Underneath a line plot shows daily 10 km-grid averaged PM_2.5_ 2.5 smoke estimates for a single site (San Diego) over the maximum possible date range. Lastly, **d)** we illustrate our estimand of interest: How wildfire pollution affects interindividual differences in intraindividual change over early adolescence (called here ‘adolescent trajectories’, ages 9 –16 years). Three subjects are plotted to illustrate interindividual differences in intra-individual change; the histogram illustrates the variance of this intraindividual change across all subjects for cortical thickness.

We start by replicating the well-established relationship between socioeconomic status (SES) and neighborhood PM_2.5_ pollution. We find a strong negative relationship (Pearson’s r = -.24, *p* < .001, BF_10_> 1000), demonstrating that people with lower SES are, on average, exposed to more neighborhood pollution. Next, we examine whether a similar association exists between SES and wildfire PM_2.5_, finding negligible/absent associations with SES for both PSE (Pearson’s r = -.02, p = .03, BF_10_= .29) and CSE (Pearson’s r = .004, p = .73, BF_10_= .03; Sup. Figure 2). This allows us to use wildfire-PMexposure to gain insights into how pollution affects neurodevelopment, thereby avoiding the confounding effects of various SES factors known to affect neurodevelopment (*12*, *26*, *30*).

Our approach leverages three strengths to ensure internal validity: *1)* the use of wildfire-specific pollution, *2)* the arbitrary jitter in the date of enrollment of children, allowing within-site variability of exposure, and *3)* longitudinal, within-subject change in neurodevelopmental outcomes. To test how wildfire PM_2.5_ (PSE/CSE) impacts cognitive and neural *trajectories* over early adolescence (defined here as ages 9—16; Figure 1d), we fit a series of mixed-effects models using four time-point data covarying for SES, neighborhood pollution, and recruitment site. PSE and CSE are tested separately, yielding estimates of their effects on 10-year-old children (i.e., intercept) and on the slope (i.e., trajectory) of development (see SI-Methods Equation 1).

The timing of within-subject neural trajectories is an area of active research (*18*, *19*), making the interpretation of cross-sectional intercept effects challenging. For instance, it is unclear if a negative or positive sign of either perinatal or childhood smoke exposure on MR modalities (at 10 years of age) indicates typical or atypical adolescent development. For this reason, we focus our interpretation on intraindividual change over early adolescence (called here ‘*trajectories’*; Figure 1d), allowing us to test our *a priori* hypothesis that prior cumulative wildfire PM_2.5_ exposure negatively impacts later cognitive and neural development. To ensure our results generalize beyond a single model specification, we conducted a series of robustness checks through alternate defensible model (re)parametrizations (‘Methods: Robustness Testing’).

### Childhood Smoke Exposure on Neural Development

To gain a comprehensive understanding of how childhood smoke exposure (CSE) impacts neural development, we focused on five structural MRI modalities (Sup. Figure 4 & 6): cortical thickness (CT), surface area (SA), subcortical volume, mean diffusivity (MD), and fractional anisotropy (FA). First, we test global neuroimaging modalities cross-sectionally. At 10-years of age (i.e., intercept), we found a significant cross-sectional CSE association with mean cortical thickness (B = .0001 mm per 10PMμg/m^3^, [*β* = .07], *p <* .001). In contrast, there was no association between CSE and total SA or total subcortical gray matter volume in 10-year-old children (*p* > .05; Sup. Table 4). For white matter diffusion measures, we found CSE to have significant cross-sectional association with average FA (B = .00006 unitless 10PMμg/m^3^, [*β* = -.04], *p<* .001) and average MD (B = .00002 mm^2^/s 10PMμg/m^3^, [*β* = .07], *p* = .004) at 10 years of age (Sup. Table 6).

We next investigated whether prior CSE influences longitudinal whole-brain developmental trajectories during early adolescence (Sup. Figure 6). We find a robust negative effect of CSE on the global neural development of all modalities, except cortical thickness (Figure 2, Sup. Figure 7, Sup. Table 4 & 6). This effect was negative, indicating diminished neural development with additional pollution exposure. We observe moderate spatial specificity (Figure 3 A & B); see supplementary materials for regional analyses (Sup. Table 10 – 14).

**Figure 2:**
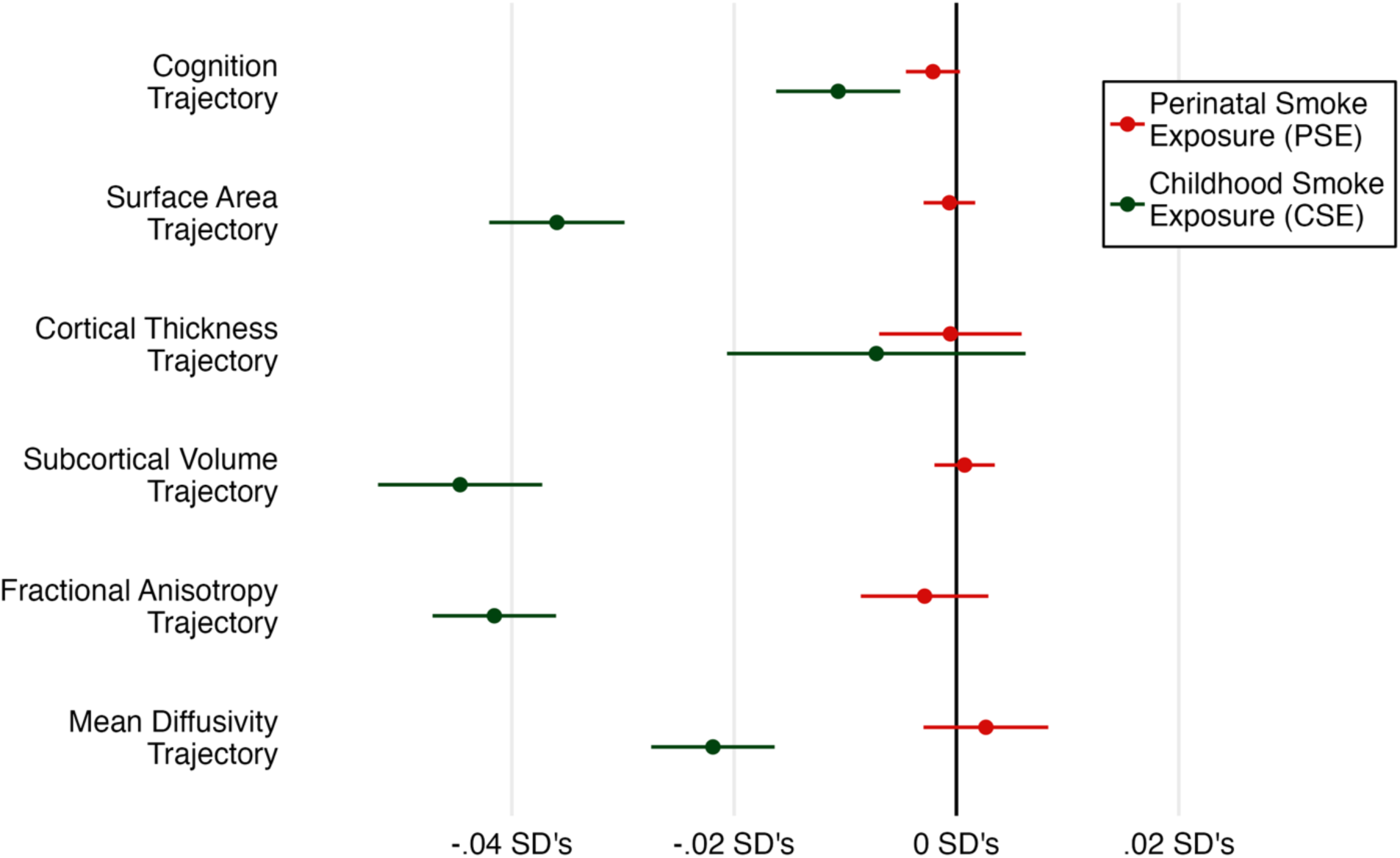
Perinatal (PSE) and childhood (CSE) smoke exposure effects on cognitive and neural adolescent trajectories (i.e., slopes). Adolescent trajectories (conceptually illustrated in Figure 1d) were defined as the interaction term between age and smoke exposure in a longitudinal linear mixed effects model (Equation 1) fit to 4 waves of data from 9 – 16 years of age. For plotting purposes, all effects are standardized (see Sup. Tables 3 – 8 for results tables). To give a sense of the magnitude of our effect sizes, we also calculate 1) weeks of divergence, and 2) the percentage of random slope variance explained for each modality (Sup. Table 9)

**Figure 3:**
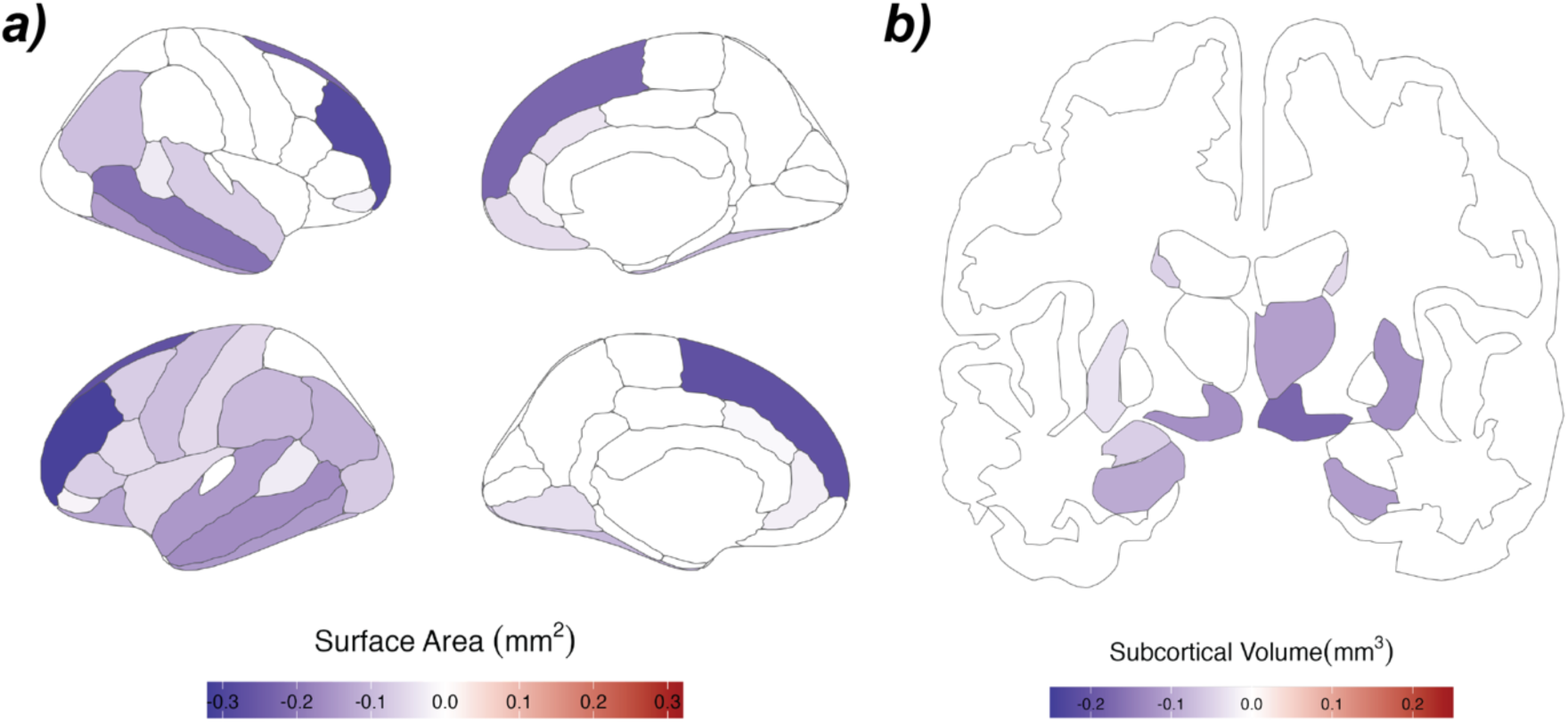
A broad negative effect from childhood smoke exposure (CSE) on adolescent trajectories (slopes) of **a)** Surface Area and **b)** Subcortical Volume. Cortical effects of surface area are predominantly in the association areas. To isolate ‘adolescent trajectories’, longitudinal mixed-effects modeling (Equation 1 in Methods) was fit to four time points from 9 to 16 years of age (see Figure 1d for a conceptual overview). Only regions remaining significant after multiple comparison correction (BY-FDR < .05) remain colored.

.Robustness testing confirmed the presence of significant negative trajectory effects (all p’s< .05 & same sign direction) for total SA (B=-6.2 mm per 10PMμg/m^3^ per year, *β*=-.04, *p<* .001) and total subcortical volume (B=-2.15 mm^3^ 10PMμg/m^3^ per year, *β*=-.04, *p<* .001). To put our findings in context, a child exposed to one standard deviation more of wildfire-PMhas roughly a year less of subcortical volumetric growth over this period of adolescence compared to a child exposed to an average level of CSE (Sup. Table 9). For white matter diffusion measures, the effect of CSE on average FA was robust (B=-.00001 unitless 10PMμg/m^3^ per year, *β*=-.04, *p<* .001), yet MD failed robustness testing (1/4 tests showed a different sign; Eq2). In summary, our whole-brain and regional results converge to show widespread negative developmental effects– across different structural neural modalities– from childhood smoke exposure (Sup. Figure 8).

### Childhood Smoke Exposure on Cognitive Performance

Analysis of cognitive scores over this period of early adolescence (9–16) showed that each year of development increased cognitive performance by 0.4 SDs (Sup. Figure 6a). We found a significant negative effect of childhood smoke exposure (CSE) on cognitive performance at 10 years of age (B=-.0012 SDs per 10 PM_2.5_ μg/m^3^, *p*=.023, [β=-.05]), and on the rate of change (i.e., slope) over adolescence (B=-0.0002 SDs per 10 PM_2.5_ μg/m^3^ per year, β=-.01, *p<*.001; Sup. Table 4). Robustness testing confirmed the presence of a negative effect of CSE on cognitive trajectories over adolescence (all *p’s*< .05 & same sign direction).

In agreement with our neural findings, additional wildfire-PMexposure negatively affected cognitive development (Figure 2). To gain an intuitive understanding of the practical effect size over these six years of adolescence, we estimate ‘weeks’ of divergent trajectories using our estimate of a year of development (i.e., main effect of age; 1 year=0.4 SDs). When we compare children with differing levels of CSE (+1σ vs -1σ), we find a developmental delay of 10-weeks in cognitive performance over these six years (-.08 SDs, Sup. Table 9 & Sup. Figure 7).

### Perinatal Smoke Exposure on Adolescent Development

Having found widespread negative neurodevelopmental effects of CSE, we moved on to test perinatal smoke exposure (PSE), as this period is characterized by enhanced neural plasticity. PSE showed a negative association with cognitive performance at 10 years of age (B=-.0026 per 10 PM_2.5_ μg/m^3^, *p* = .014, [*β*=-.02]), yet no effect on change over adolescence (B=-0.0001 per 10 PM_2.5_ μg/m^3^, *β*=-.00, *p* = .088; Sup. Table 3).

Cross-sectionally, only average cortical thickness at 10-years of age was significantly associated with PSE (Sup. Table 5&7). Similar to our cognitive performance results, there was no significant relationship between PSE and any whole-brain adolescent structural neural trajectories. When testing regional trajectories across our five MRI modalities, only a single fiber track (FA of the left striatal inferior frontal cortex) survived multiple comparison correction (Sup. Tables 10 - 14). This broad absence of any perinatal effect could be due to a variety of factors, such as protective factors of the womb, a more limited range of exposure, or the extended period of time between exposure and adolescent development.

## Discussion

Leveraging a large neurodevelopmental cohort and a natural experimental framework to decouple pollution from socioeconomic factors, we isolate the effects of wildfire PM_2.5_ on neural and cognitive development. Doing so, we find broad negative effects of wildfire pollution during childhood on subsequent neural and cognitive development over adolescence. Within-subject neurodevelopmental effects are present across every structural neuroimaging modality studied (except cortical thickness), and are robust to different model specifications. Our findings add causal weight to prior human neuroimaging work using more confounded sources of PM_2.5_ pollution, such as neighborhood pollution (*24*), and add evidence to the urgent public health concerns of particulate matter pollution on childhood development (*3*, *11*).

In addition to our neural results, we find wildfire-PMexposure negatively impacts cognitive development – showing a 10-week developmental delay when comparing children with differing CSE exposure levels (+1σ vs -1σ). This result is equivalent to a medium-sized educational intervention (*51*) and consistent with prior causal inference work showing that PM_2.5_ negatively affects math and language test scores cross-sectionally (*39*, *52*).

The widespread neural effect we find is consistent with mechanistic accounts from animal models showing PM_2.5_ entering the bloodstream, crossing the blood-brain barrier, causing neuroinflammation and oxidative stress, altered neural and glial cell morphology, and impaired blood–brain barrier integrity (*3*, *4*, *11*) – these mechanisms could either slow neurodevelopment and/or cause neurodegeneration. Although we generally see widespread and robust effects, they were not identical across all modalities or brain regions. Specifically, cortical thickness and mean diffusivity do not demonstrate broad negative developmental effects, despite not differing in properties such as reliability (which would affect power; Sup. Fig. 4). A more plausible interpretation is that distinct neurobiological components (e.g., myelination, dendritic arborization), which our neural metrics are proxies for, are differentially vulnerable to the negative effects of wildfire pollution (*11*, *31*). Alternatively, differential developmental curves (*14*, *18*, *30*) across structural modalities would likely lead to varying windows of sensitivity and malleability to CNS toxicity. We find some support for this within-modality, for instance, the effect of CSE on regional surface area development is present predominantly over the association areas, indicating pollution affects later-developing, cognitively relevant brain regions (*15*). Alternative mechanisms adversely impacting neural and cognitive development (e.g., the long-term psychological outcomes of wildfires) are also possible, although we note that children with high wildfire PM_2.5_ 2.5 exposure levels are not necessarily those within the direct proximity of a wildfire (due to prevailing westerly winds). More work is needed to determine which of these factors is the predominant driver of the results we see.

In addition to childhood smoke exposure (CSE), we also tested perinatal exposure (PSE), inspired by theoretical accounts (*14*, *17*, *48*) and empirical findings (*49*, *50*), which highlight the unique and particularly strong impacts of environmental exposure during this period. However, in contrast to the robust CSE effects, we did not find any replicable neurodevelopmental effects from additional PSE. Several explanations are possible. First, the lack of effect may be genuine– for instance, if either biological (e.g., placental filtering) or behavioral (e.g., maternal avoidance behaviors) factors limit exposure. Second, due to the design of our study, PSE may cause immediate adverse effects on early brain development, which are remediated through neuroplasticity and no longer present during adolescence. Notably, we do find weak cross-sectional evidence of lasting PSE exposure on cortical thickness and cognition. Finally, the range of perinatal PM_2.5_ exposure is lower than during childhood, making an effect (if it exists) less easily detectable even in such a large sample.

In summary, we report widespread and robust cognitive and neural developmental consequences from additional wildfire-PMexposure in over 9000 children from the United States. Our findings are of considerable translational importance; almost the entire world (99%) lives in areas with PM_2.5_ pollution above WHO health guidelines (*2*). Crucially, air pollution regulation commonly excludes wildfire pollution (e.g., the Clean Air Act), despite this type of pollution representing a steadily increasing problem within and beyond the United States (*41*). Given the lifelong impacts of neurodevelopmental trajectories in childhood and adolescence, it is paramount that we better understand and, ultimately, devise ways to mitigate the harmful effects of PM_2.5_ and wildfire pollution.

## Data Availability

Data used in the preparation of this article were obtained from the Adolescent Brain Cognitive Developmen (ABCD) Study, held in the NIH Brain Development Cohorts Data Sharing Platform. The ABCD Study is supported by the National Institutes of Health and additional federal partners under award numbers: U01DA041048, U01DA050989, U01DA051016, U01DA041022, U01DA051018, U01DA051037, U01DA050987, U01DA041174, U01DA041106, U01DA041117, U01DA041028, U01DA041134, U01DA050988, U01DA051039, U01DA041156, U01DA041025, U01DA041120, U01DA051038, U01DA041148, U01DA041093, U01DA041089, U24DA041123, U24DA041147, NIEHS R01-ES032295 and R01-ES031074. ABCD Consortium investigators designed and implemented the study and/or provided data but did not necessarily participate in the analysis or writing of this report. This manuscript reflects the views of the authors and may not reflect the opinions or views of the NIH or ABCD Consortium investigators.

https://docs.abcdstudy.org/latest/

## Statistical Analysis Plan

To determine the impact of *prior* wildfire PM_2.5_ pollution on cognitive and neural levels (e.g., intercept, at 10 years of age) and the change (e.g., slope) over six years of development, we combined two open-access data sets; *1)* high-resolution daily local-level PM_2.5_ wildfire smoke estimates (v 1.0; (*1*)) (*2*) and *2)* behavioral and neural data from the Adolescent Brain Cognitive Development (ABCD v 6.0; (*3*)) consortium (*4–6*). Our paper focuses on change (called *‘trajectories’*) over adolescence, yet all intercept effects are reported in the supplement.

The ABCD consortium is an ongoing multi-center study that recruited 9 – 11-year-old children (starting N = 11,868; total N = 9,806) and is longitudinally following them in two-year increments for ten years. Our analysis focuses on the first six years with four approximately equally 2-year spaced waves (ABCD v6.0). As the ABCD consortium includes twins and siblings, we sampled the individual with the most follow-up attendance (if equal random sampling was used), resulting in a sample of 9806 children in wave 1. Ethics was reviewed and approved by the central institutional review board at the University of California, San Diego. Written parental informed consent, along with child assent, was obtained for all participants (*4*).

### Constructing wildfire smoke PM_2.5_ estimates

We harnessed well-validated and reliable daily local-level PM_2.5_ wildfire smoke estimates for the contiguous United States (v1.0) (*2*) to measure a child’s cumulative exposure to wildfire smoke. We first masked a geographical area equal to the ABCD 50-mile subject recruitment catchment criteria (*4*) centered on each of the 22 ABCD recruitment sites (Figures 1 & 4). The ABCD consortium probability sampled schools from these sites; the coverage of these sites represents ∼20% of the 9-11-year-old population in the United States (see (*4*)).

**Figure 4:**
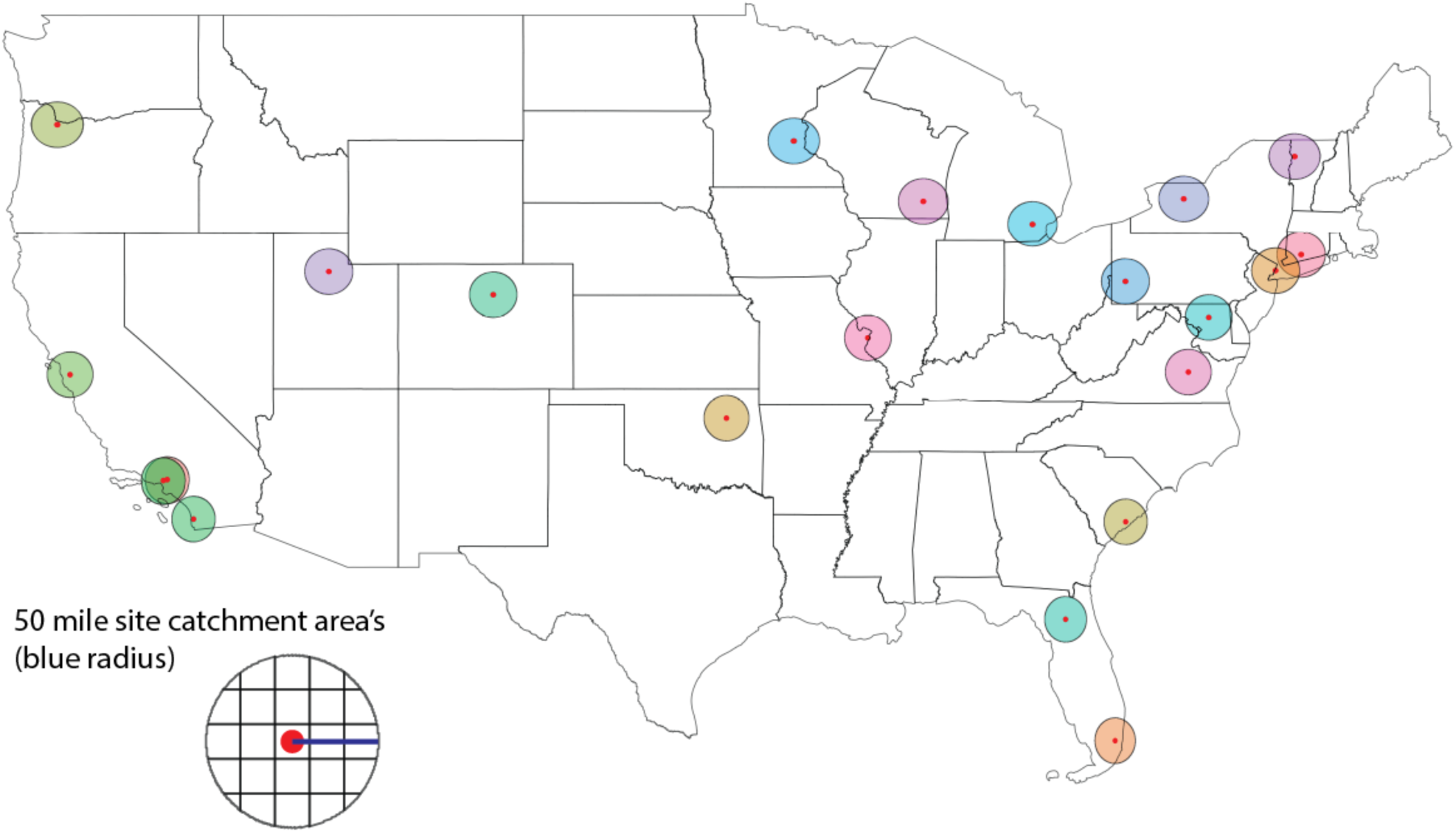
An overview of the 22 ABCD recruitment sites and their 50-mile catchment areas (represented as black circles). The red dots reflect a much smaller (only 3.7%) 10km radius catchment area used for our first spaBal sensiBvity analysis; we also illustrate the 5-by-5 grid used in the 3^rd^ spaBal sensiBvity analysis. This series of analyses found no evidence for within-site disBnct geographical variance for either of our smoke exposure measures (PSE/CSE).

We then calculated the daily pollution per site by averaging the 10km grid wildfire PM_2.5_ estimates inside each catchment area. In a series of spatial analyses, we found wildfire PM_2.5_ pollution estimates do not show local-level (e.g., 10km) geographic specificity, limiting city-level spatial variation (see Methods section ‘*Within-site Spatial Stability’*) for cumulative measures. We have the rather simplistic assumption of residential stability within a site, as in children can move, yet ideally not between sites. Over the 6 years of longitudinal sampling, we find 1.8% change their site at least once – assuming site-movement rates are equal between 9–11 and birth until 10; this would mean ∼3% of children moved sites during our exposure measures. This would most likely add noise to our exposure measure, rather than any sort of systemic bias. Moreover, this small percentage makes it highly unlikely that the assumption of residential stability would affect our findings.

Each child’s date of birth was used to build individual-specific smoke exposure measures. In ABCD release 6.0, the date of birth was anonymized in two buckets per month (first half & last half). For instance, if a child was born on the 1^st^ of June 2008, they are indistinguishable from a child born on the 10^th^ of June 2008. Since there are two buckets per month, a range of around 15 days is anonymized. We wanted to ensure we did not miss PM_2.5_ exposure; therefore, we set every child in a 15-day bucket to the oldest age possible within this bucket (at the cost of overestimating exposure).

Due to the recruitment characteristics of ABCD, an early-born child will not necessarily have more exposure than a later-born child. Age is not deterministically related to exposure, as the consortium recruited a window of children (9 – 11 years of age) over a long baseline recruitment phase (i.e., range of 26 months between the first and last baseline visit), resulting in four years of age range between the youngest and oldest-born child (49 months). This variance contributed to substantially different exposure levels between children.

Childhood Smoke Exposure (CSE) was defined as cumulative PM_2.5_ μg/m^3^ wildfire exposure from 3 months of age up until the first behavioral timepoint of ABCD (Figure 1a). This resulted in a CSE measure for all eligible children, bar one, whose date of birth (plus 3 months) fell outside of the range of smoke exposure measures (CSE; N = 9805, mean = 1145 PM_2.5_ μg/m^3^, SD = 455 PM_2.5_ μg/m^3^). This limit is due to the National Oceanic and Atmospheric Administration Hazard Mapping System smoke plume data starting in 2006 (*2*). We also constructed a perinatal smoke exposure (PSE) measure by summing cumulative wildfire exposure in the first 3 months of life and 9 months prior (i.e., conception as is established practice (*7*)). We only included children with this entire period from 2006 onwards (due to missing NASA smoke plume data), resulting in the exclusion of 9.4% of eligible children (N = 926) with gestation periods in 2005. We had PSE data for 8880 children (mean = 140 PM_2.5_ μg/m^3^, SD = 94 PM_2.5_ μg/m^3^). Childhood and perinatal smoke exposure were weakly correlated (Pearson’s r = 0.07; Sup. Figure 2).

### Wildfire exposure and its relationship with Socioeconomic Status

Prior work (*8*, *9*) (including our findings) has shown wildfire PM_2.5_ pollution to be unrelated to socioeconomic status (SES). Yet, avoidance behaviors (e.g., staying inside) and infrastructure differences (e.g., home air filtration) can lead to individual differences in attained exposure, which most likely follow an SES gradient (*10*). Prior work attempting to explicitly show this for wildfire did not show such an association (*8*), although SES-gradient avoidance behaviors have been shown for other pollutants (*11–13*).

Crucially, even in the case of avoidance behaviors, it is highly improbable to be a threat to the exogeneity of our statistical inferences (i.e., Type I error, false positive), as differential-SES avoidance behaviors would lead us to *underestimate* our effects (Type II error; biased towards the null). The variation in our exposure measure is unrelated to SES (assumption: no causal path from SES -> to IV variables [PSE/CSE]), thereby uncoupling our IV (wildfire pollution measured as PSE/CSE) from endogenous SES variance. As with an intent-to-treat analysis, attained exposure differences can still occur without introducing Type I bias (i.e., false positives). Conversely, if we measured attained wildfire pollution exposure through in-house sensors (akin to an analysis by treatment received) and pollution followed a differential SES gradient (whereby low-SES individuals attain more wildfire pollution) in the case of a significant effect, it would be unclear if this is a causal effect or a false-positive (Type I error). Similar to neighborhood pollution, our exposure variable would be inherently linked to SES – thereby, making it unclear to what extent our estimate reflects a true causal effect or is driven by one of the many SES-related factors affecting neurodevelopment. This statistical point does not negate *nor is it in opposition* to the existence of avoidance behaviors, causing differing levels of wildfire PM_2.5_ pollution exposure across SES strata. While it is likely the case, more work is needed to solidify the existence of this phenomenon.

### Behavioral and Demographic Measures

To estimate the development of cognitive ability during childhood and early adolescence, we leveraged the richness of cognitive tests available in ABCD. Confirmatory factor analysis (CFA) was used to combine four cognitive tests (Picture Vocabulary, Picture Sequence Memory, Oral Reading Recognition, and Working Memory n-back correct response rate) (*14*, *15*) into a latent *cognitive performance* factor for each time point (see Sup. Figure 5 for a correlation plot). Doing so increases power by removing measurement error (rather than focusing on each cognitive task separately). In line with best practice (*16*), all models were specified with correlated error variance within task across time (Sup. Figure 3). The lavaan R package (v. 0.6.19) (*17*) was used with full-information maximum-likelihood estimation and a robust maximum-likelihood estimator with a Yuan–Bentler-scaled test statistic. In line with best practice, missing or incomplete test data were dealt with using Full Information Maximum Likelihood estimation(*18*). This approach leverages all available data, increases power, and decreases bias compared to simply omitting individuals with partial data (*19*). We determined adequate (CFI > .9; RMSEA < .08) and good (CFI > .95; RMSEA < .05) fit using the CFI and RMSEA cutoffs (*20*).

We then tested for longitudinal measurement invariance to ensure the latent factors represent the same concept throughout time – a *prerequisite* for any change-based analysis using summary measures. Briefly, a series of model comparisons, in ascending order of strictness, is run to determine longitudinal measurement invariance (see (*21*)). Due to the large sample size of ABCD, we tested invariance via a change of ≥ −.010 in CFI, supplemented by a change of ≥ .015 in RMSEA (*22*). We did not test strict measurement invariance (error variances fixed through time) as this was not necessary for our statistical inferences. A strong measurement invariance longitudinal CFA fit the data well (χ^2^=1941.6, df =112, p <0.0001, RMSEA = .044, CFI = .962) and passed measurement invariance (Sup Table 1). Next, we extracted factor scores from this model as our measure of cognitive performance. This measure was standardized using the first timepoint (mean_T1=0; SD_T1=1).

Neighborhood PM_2.5_ was included as part of the ABCD study, and is defined as the average annual PM_2.5_ exposure at the subject’s residential address in 2016 (*23*). Part of this measure includes wildfire PM_2.5_ from 2016 and should be seen as overcontrolling (increasing the chance of a type II error to avoid a type I error; ergo, limiting false positives). Socioeconomic status (SES) was defined as the first principal component from a probabilistic PCA of total household income, highest parental education, and neighborhood quality. We did not use the consortium’s SES composite, as our approach allows for missing data. SES was standardized with a mean of zero and a standard deviation of one (for more details, see (*24*)), with higher scores reflecting more parental income/education and less neighborhood deprivation. A small minority of children (1.8%, N = 181) changed their ABCD site at least once throughout the four follow-up waves; for those children, we control for the first ABCD site recorded. When we report correlations between PM_2.5_ measures and SES, using the cor.test function from the ‘stats’ package for frequentist (Pearson’s R) results and correlation BF_10_ with default priors from the ‘BayesFactor’ package (*25*) to report Bayes Factors. BF_10_ denotes Bayes Factor values providing evidence for the alternative hypothesis.

### Neuroimaging Procedures

Neuroimaging was performed following standardized protocols, and preprocessing utilized a uniform pipeline at a single location, which has been comprehensively outlined previously (see (*5*, *6*). We used ABCD release 6.0, which has 4 waves of neuroimaging data spaced 2-years apart (avg. ages 9 – 16) – this length of time helps increase longitudinal reliability (*26*). Briefly, data were collected using a 32-channel head coil in 3T scanners across 22 sites following a standardized protocol across three different scanner manufacturers (GE/Phillips/Siemens). T1-weighted and T2-weighted structural images were collected. Preprocessing consisted of gradient nonlinearity distortion correction, mutual information registration, intensity non-uniformity correction, and resampling with 1 mm isotropic voxels into rigid alignment (for more details, see (*5*)). For cortical surface reconstruction, Freesurfer was used, and we analyzed subcortical volume from structures labeled with atlas-based segmentation (*27*), along with cortical thickness and surface area from the Desikan-Killiany cortical atlas (*28*). The diffusion MR acquisition (1.7mm isotropic) used multiband EPI with a slice acceleration factor of three and included 96 diffusion directions, seven b=0 frames, and four b-values. Preprocessing and DTI analysis used conventional methods (see (*5*)). We use full shell DTI and only analyze mean diffusivity (MD) and fractional anisotropy (FA) using regions of interest from AtlasTrack (*29*). Voxels which are primarily CSF or gray matter are excluded from analysis; we test all 34 ROIs from the DK atlas. Statistical analysis is described in detail under the section ‘inferential approach’.

The ABCD consortium used automated and manual methods to assess data quality (*5*), in addition to manual review of all images by trained professionals to detect artifacts or abnormalities. We only include neuroimaging data that passed all quality control criteria for that relevant domain (i.e., structural or DTI QC). To further ensure that no preprocessing errors affected our analysis, we removed observations that were 5 standard deviations above or below the mean on all neuroimaging measures. This affected very few datapoints; for instance, the global measure most affected – mean diffusivity – had only 7 observations out of 22898 removed (0.03%).

### Inferential Approach

We use longitudinal mixed effects modeling to examine how prior wildfire PM_2.5_ smoke exposure impacted cognitive and neural levels (i.e., intercept) at 10 years of age and change over six years of adolescence (i.e., slopes/trajectories). Our primary model included a random slope for our measure of time (i.e., age) and a random intercept per subject nested within the ABCD site (see Equation 1 in Wilkinson notation). As we have two early life wildfire PM_2.5_ exposure measures (perinatal = PSE, childhood = CSE), the model was fit once with CSE and again with PSE as the independent variables of interest. Age was centered to reflect the average age at the first timepoint (10 years; 120 months) to help with the interpretation of the intercept.

For each cognitive and neural outcome (*Y in* *equation 1* *below*) we examined the effect of perinatal and childhood smoke exposure separately (PSE/CSE; see section ‘wildfire smoke PM_2.5_ estimates’) on the initial level at 10 years old (intercept; illustrated by the main effect in blue font) and the change over four years of development in children (slope; illustrated by the interaction term in magenta font). To properly control our slope estimate, we included all interactions between age and our control variables (matrix Z) with age (i.e., interaction-controls) (*30*). All models included socioeconomic status (SES) and ABCD neighborhood PM_2.5_ exposure as covariates of no interest; neuroimaging outcomes included dummy control variables for scanner type and sex.

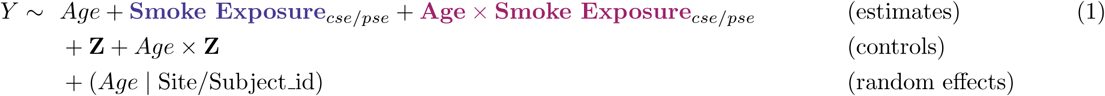

Models were fit using maximum likelihood estimation from the R package lme4 (v. 1.1.36)(*31*), and p-values (alpha = .05) were computed using Satterthwaite’s degrees of freedom method with the lmerTest package (v. 3.1.3) (*32*). All parameter estimates are reported in their raw units, for standardized beta’s we scale all model terms (mean = 0; sd =1). Standardizing affects the location of the intercept (e.g., moving it from 10 years of Age to the mean age in the sample); in models with interactions, this, in turn, affects the p-values. This does not affect the interaction term; we report p-values from the unstandardized model throughout the manuscript. For added clarity when reporting standardized intercept effects, we use square brackets around the standardized result. Our regional neuroimaging outcomes are false discovery rate (Benyamini-Yekutieli – FDR with dependency) corrected using the number of regions per modality (e.g., 68 regions for CT/SA), with a q value below 0.05 considered significant (abbreviated as *p*_FDR_).

### Statistical Robustness Testing for Global Effects (Equations 2 – 5)

To ensure transparency and replicability, we tested a series of theoretically defensible alternative model parametrizations to ensure our results are robust to specific model specification (a practice common in economics) (*33*). This allows us to ensure that our inferences are not contingent on small methodological choices – If an estimate is substantially different across parametrizations, we consider it not robust, and limited to a narrow model specification (in our case, Equation 1). To do so, for models with *significant* global slope effects (i.e., interaction term in magenta font), we compare the correspondence between the sign and significance of our alternative model parametrizations (Equations 2 – 5). We do not perform robustness testing on the global cross-sectional effect of Smoke Exposure at 10-years of age (intercept; illustrated by the main effect in blue font) as main effects in the presence of an interaction by design will change sign and significance due to centering (*34*). A result is deemed ‘robust’ if the estimate remains significant and the sign is in the same direction across all tests (magenta term in equations 2 – 5).

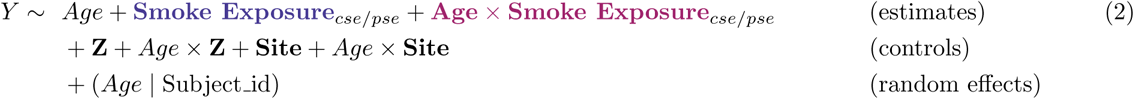

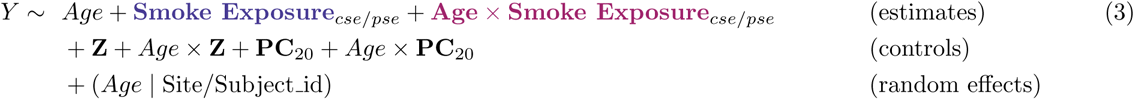

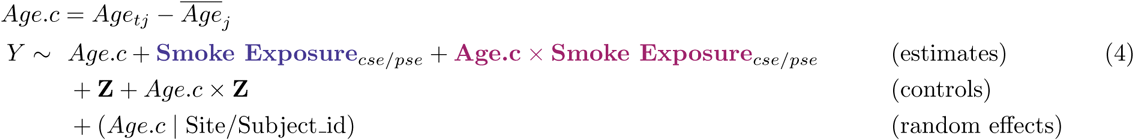

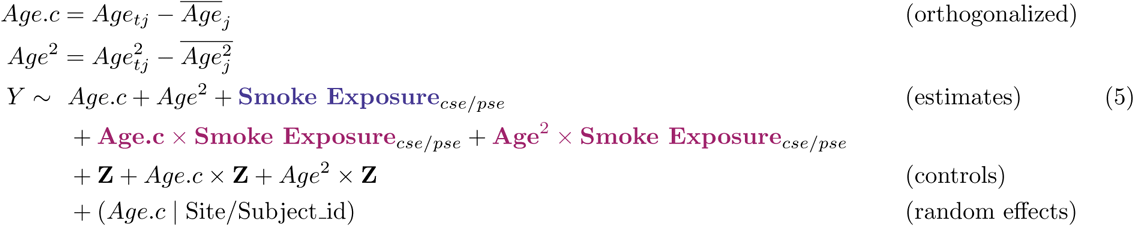

For instance, our main model codes site as a random intercept (as recommended by the ABCD consortium (*35*)), yet this assumes ‘sites’ to be a random sample from the same underlying population, which is not the case in the ABCD consortium (*4*). In Equation 2, we solve this limitation by replacing our site random intercept with a matrix of dummy-coded fixed effects in addition to interaction controls (e.g., Site1*Age)(*30*), treating each site as an independent variable. To further ensure generalizability across (overlapping) populations, Equation 3 adds matrix PC_20_, representing 20 genetic principal components, controlling for the fixed and interaction effects related to genetic ancestry^1^.

The ABCD consortium followed up individuals in 2-year increments for four waves (release 6.0) spaced in (approximately) 2-year age buckets. In this type of design, ‘age’ is coding for time (as there is only one cohort); nevertheless, to *ensure* we are interpreting within-individual change, we conducted a within-person age-centered model (Equation 4), including only individuals with at least 2 timepoints.

In Equation 5, we extend this to the quadratic model for individuals with at least 3 timepoints. Crucially, we use the ‘poly’ function in R to construct a 2^nd^ degree orthogonal polynomial of age and then mean-center these two terms (rather than the reverse; mean-centering and constructing polynomial terms). This minor choice *dramatically* changes the interpretation of ‘age’ from one of absolute (chronological age) to relative age (biological age). Our (Equation 5) approach is preferred when focusing on average within-individual age patterns with quadratic effects (*36*). We use model comparison to test if the additional quadratic term improves model fit before running this additional robustness test. We find this to be the case in all modalities (Sup. Table 2) except average cortical thickness and average fractional anisotropy. We find our global MR modalities to have a linear pattern over adolescence (10 – 16 yrs), see Sup. Figure 6 for linear [black line] and quadratic [red line] fits overlayed and Sup. Table 2 for model comparison. This series of robustness tests confirms the adequacy of Equation 1 as our main model of inference.

### Missing Data

Missing data came from two sources: *1)* missing wildfire smoke estimates necessary to construct exposure measures, and *2)* missing ABCD consortium measures. The first source was due to smoke pollution estimates starting in 2006, as this is the first year of complete NASA smoke plume data (*2*). Fortunately, most children were born after 2005, so we only need to exclude one child for our CSE measure. Yet, 9.4% of eligible children (N = 926) had gestation periods in 2005, resulting in missing PSE measures for these children. This first source of missingness is deterministic based on birthdate; therefore, it most likely does not present issues to our estimate of interest.

ABCD made significant efforts to recruit an initially representative sample of the United States (*4*, *37*). Wave 4, compared to the other waves, has fewer subjects with imaging data than the other 3 waves. A large part of the missingness from the 4^th^ wave is due to ABCD v6 being a partial release. Another source of missing data is due to observable characteristics (MAR) or reasons unknown to us (MNAR). For instance, socioeconomic status (SES) is an observable characteristic that is known to impact the probability of *1)* initial enrollment and *2)* follow-up attendance in large consortium studies (*38*). Our modeling framework, using FIML, will yield unbiased estimates if missingness varies as a function of characteristics also included in the model.

We employ a variety of approaches to minimize the impacts of differential missingness. First, we use a probabilistic PCA to estimate each child’s SES; unlike standard principal components methods, this allows us to include subjects with partially missing data (*24*). Due to this choice, for the initial sample of eligible children (N = 9806) none are missing baseline socioeconomic status data. All models include terms for SES and neighborhood PM_2.5_, in turn, accounting for MAR missingness based on these variables. Lastly, we used full information maximum likelihood for missing cognitive performance task data.

### Within-site Spatial Stability of wildfire smoke PM_2.5_ estimates

Next, we examine the within-site spatial stability of smoke exposure, inspired by the geographical differences in smoke exposure across the United States (*39*). What is less clear is to what extent local (i.e., within-city) differences in wildfire smoke exposure contribute to varying cumulative exposure profiles. In other words, is it likely that two individuals within the same ABCD site experience are exposed to substantially different levels of *cumulative* prenatal and childhood wildfire smoke exposure? To examine this question, we test the within-site stability of PSE and CSE. On average, each site’s 50-mile catchment area included 259 10km grids of unique wildfire PM_2.5_ smoke pollution predictions (range: 169 – 342).

If our exposure focused on individuals near discrete wildfire events, this level of coarseness would be ill-suited for mapping exposure profiles, as in the near vicinity of wildfires, there are distinct spatial gradients of pollution. Yet as wildfire pollution spreads, the gradient of exposure does as well – wildfires in California contribute to pollution in states as far away as Maine. Ultimately, it is an empirical question: *how much could we have possibly gained from within-site geographical variance with more precise (non-anonymized) geographical information, such as a child’s residential address?*

If site-specific geographical differences in PM_2.5_ exposure on the same day were a primary driver of pollution exposure, it would have impacted the power of our design, increasing our chances of null findings (Type II error), and likely causing an underestimate of the true effect. To accomplish this, we took three complementary strategies in which we estimated PSE and CSE exposure measures per individual: *1)* in a much smaller catchment area centered on the site, *2)* using random, within-site single grid sampling, and *3)* by dividing the larger catchment into 25 ‘neighborhood’ grids.

First, we constructed a much smaller 10km site catchment area around each imaging center. On average, these smaller catchment areas included only nine 10km grids of smoke pollution estimates (range: 8 – 12). Despite the substantially smaller size (∼3.7% of the area; red circle in Figure 4), both PSE & CSE person-specific cumulative exposure estimates correlated extremely highly (Pearson’s r = .98) with estimates using 50-mile catchment areas. This is impressive, especially considering the two estimates correlate very weakly with each other (Pearson’s R = .07). Next, we calculated PSE and CSE cumulative exposure estimates by randomly sampling a *single* 10km wildfire grid with daily estimates within that individual’s 50-mile catchment area. This approach *maximizes* geographical within-site variance at the cost of increasing error variance by relying on a single estimate per day. Repeating this 100 times, we found the average correlation to be very high for both PSE (avg. Pearson’s r = .85) and CSE (avg. Pearson’s r = .81).

Both approaches neglect important aspects that would be considered when calculating a pollution exposure measure with an exact residential address. First, centering by imaging site is most likely not representative of true within-site variance (as wildfires are more prevalent near the peripheries of cities). Moreover, values from the smaller catchment area are part of the larger (biasing the correlation upwards, although to a minor extent). Random sampling is also less than ideal as it forgoes the stability gained from averaging across multiple grids on the same day (lessening any potential estimation error). Therefore, we conducted a third analysis where we divided 50-mile catchments into a 5-by-5 grid square (Figure 4) – allowing us to have larger patches of geographically neighboring wildfire estimates within a site.

Since we divided a *circular* catchment into a 5-by-5 *square* grid, some areas (for instance, all the corners) included less than ten 10km estimates of smoke pollution and were therefore excluded. This made the maximum number of possible grid ‘neighborhoods’ 21 (25 - 4 corners), yet each site had varying amounts (mean = 19.7). Using the remaining grids, for individuals from that specific site, we calculated PSE and CSE exposure measures per ‘neighborhood’ grid and correlated them. This resulted in exceptionally high correlations between PSE (mean Pearson’s R = .94) and CSE (mean Pearson’s R = .97) exposure measures of geographically distinct neighborhood grids – again, indicating within-site variance of cumulative smoke exposure to be minimal.

All three within-site geographical analyses converge to indicate limited *within-site* spatial specificity for our cumulative smoke exposure measures. The meaningful variability in our cumulative wildfire PM_2.5_ exposure estimates is primarily due to differences in children’s date-of-birth, first visit date, and site-wide geographical effects. Moreover, a lack of within-site spatial specificity adds to the strength of wildfire pollution as an instrument for the purposes of a natural experimental design, as if it is equal within-site, it will also be uncorrelated with other within-site variables (e.g., within-city SES).

### Code and Data Availability

*All code is publicly available (*https://github.com/njudd/neuralfire*). Wildfire pollution estimates are publicly available here* https://dataverse.harvard.edu/citation?persistentId=doi:10.7910/DVN/DJVMTV, ABCD data is also publically available yet must be applied for see here (https://nbdc-datashare.lassoinformatics.com/*) for more information*.

### Funding & Acknowledgments

Nicholas Judd is supported with a Jacobs fellowship from the Jacobs Foundation, along with the Pro Futura Scientia Fellowship from Riksbanken Jubileumsfond (PF25-0005). Rogier Kievit is supported by a Hypatia fellowship from the RadboudUMC.

Data used in the preparation of this article were obtained from the Adolescent Brain Cognitive Development™ (ABCD) Study, held in the NIH Brain Development Cohorts Data Sharing Platform. The ABCD Study® is supported by the National Institutes of Health and additional federal partners under award numbers: U01DA041048, U01DA050989, U01DA051016, U01DA041022, U01DA051018, U01DA051037, U01DA050987, U01DA041174, U01DA041106, U01DA041117, U01DA041028, U01DA041134, U01DA050988, U01DA051039, U01DA041156, U01DA041025, U01DA041120, U01DA051038, U01DA041148, U01DA041093, U01DA041089, U24DA041123, U24DA041147, NIEHS R01- ES032295 and R01-ES031074. ABCD Consortium investigators designed and implemented the study and/or provided data but did not necessarily participate in the analysis or writing of this report. This manuscript reflects the views of the authors and may not reflect the opinions or views of the NIH or ABCD Consortium investigators.

We would like to acknowledge Dr. Sebastian Ghormi, Dr. Léa Michel, and Dr. Jessica Schaaf.

### Ethical Statement

The Adolescent Brain Cognitive Development (ABCD) study, was reviewed and approved by the central institutional review board at the University of California, San Diego; in addition to local ethics board review at each participating site. Written parental informed consent, along with child assent, was obtained for all participants.

---

**Sup. Figure 1:**
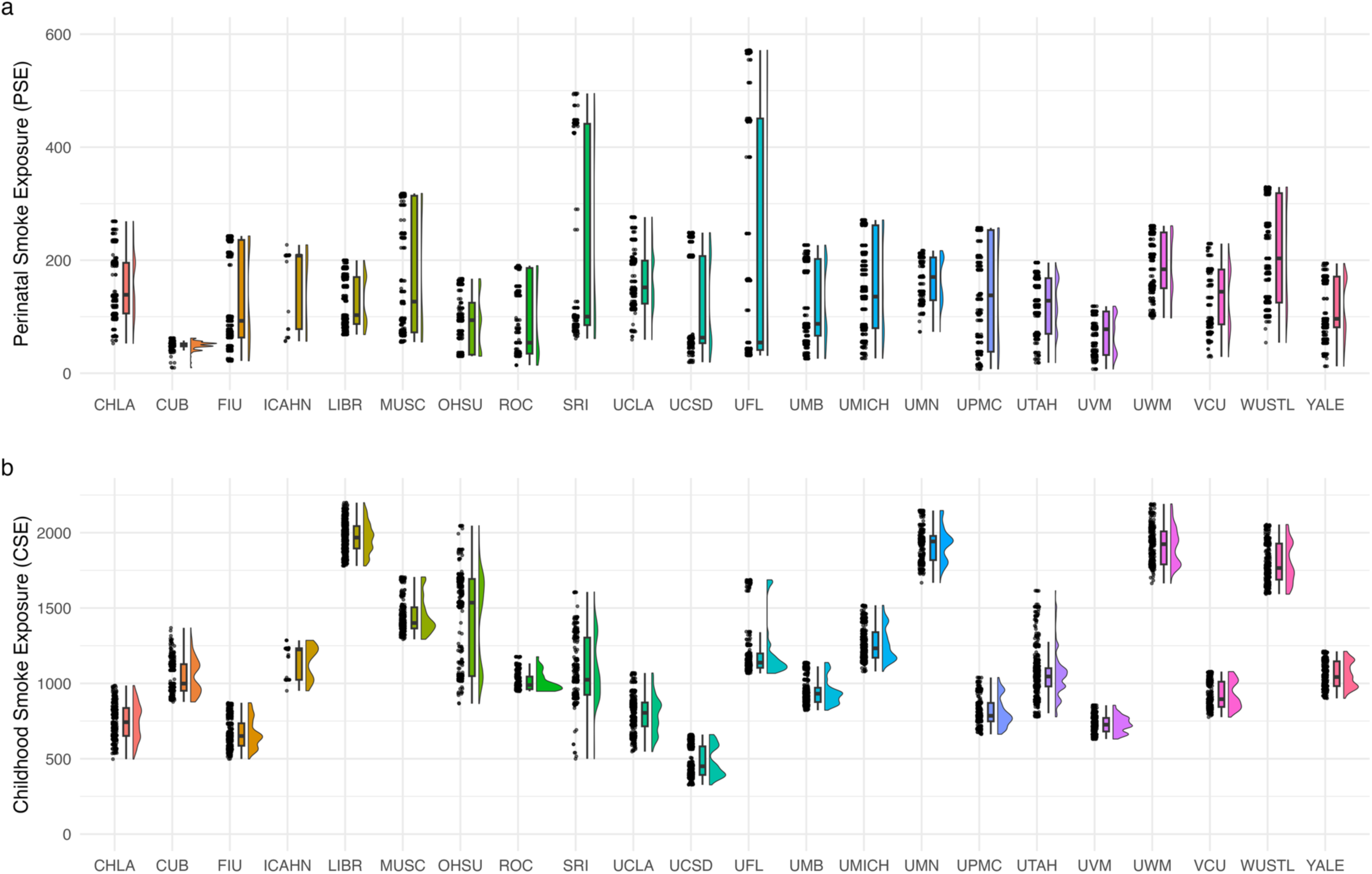
Raincloud plots illustrating substantial within and between-site variance for perinatal smoke exposure (PSE) and childhood smoke exposure (CSE). Between-site variance reflects both true exposure differences and statistical dependency (through unobserved factors making individuals more similar when sampled from the same site; breaking independence), therefore, all models correct for recruitment site. We also include fixed effects site correction as a robustness test (Equation 2).

**Sup. Figure 2:**
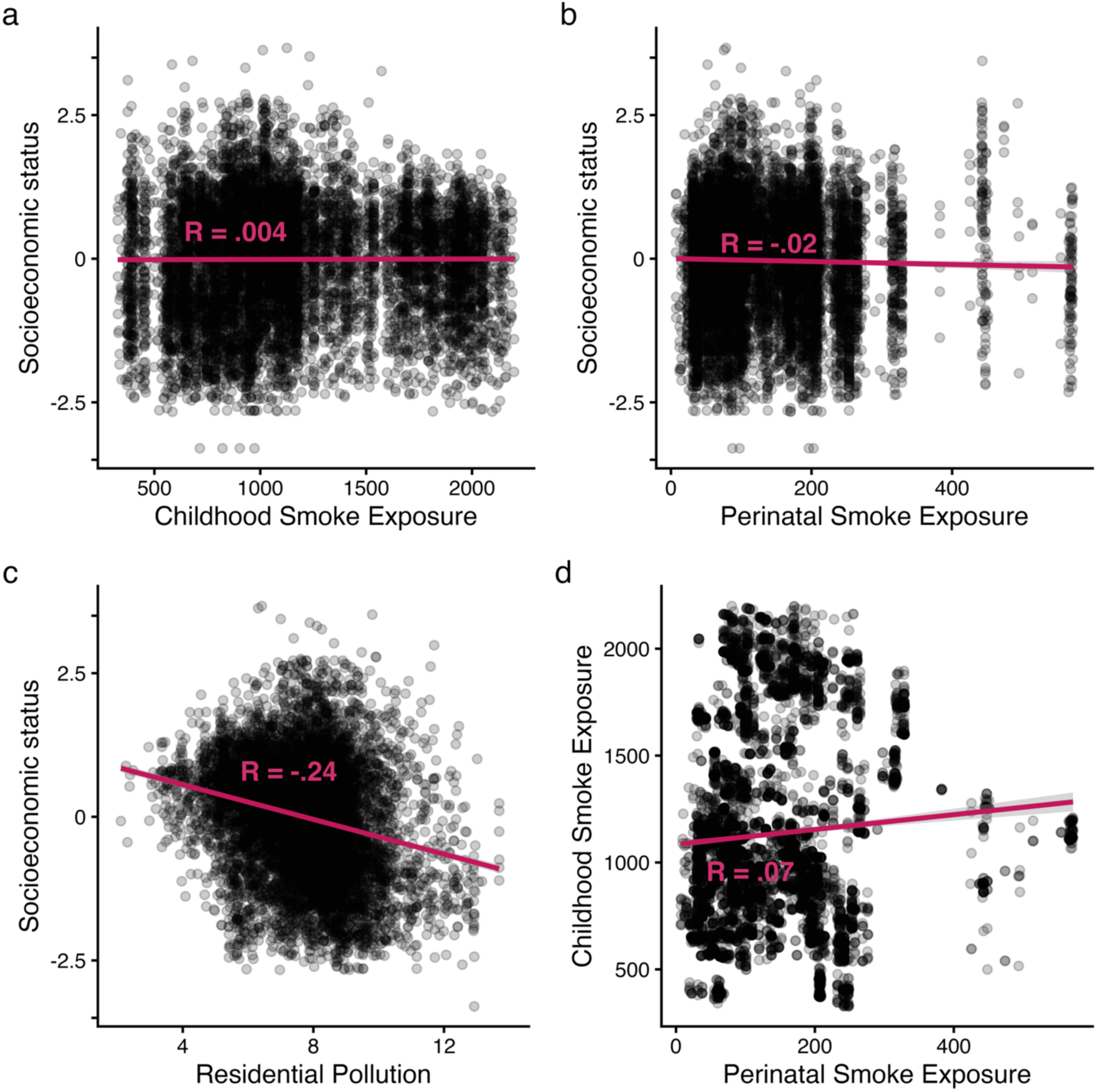
Correlation plots (Pearson’s r) between socioeconomic status and a) Childhood Smoke Exposure, b) Perinatal Smoke Exposure, and c) Neighborhood Pollution – illustrating the lack of a strong correlation between our smoke exposure measures and socioeconomic status. Lastly, d) illustrates the correlation between our two (non-overlapping) smoke exposure measures perinatal (cumulative exposure from conception until 3 months of age) and childhood (cumulative exposure from 3 months of age until the first testing occasion ∼10 years of age).

**Sup. Figure 3:**
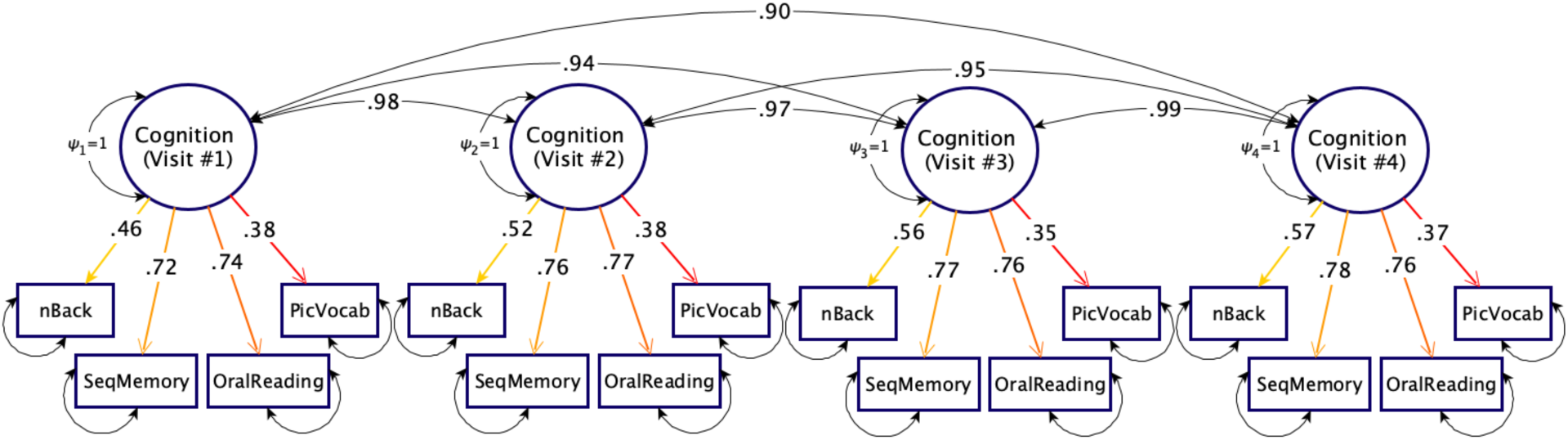
A strong measurement invariant CFA fit to four cognitive tasks across the four ABCD timepoints (Visit #1: age 10; Visit #2: age 12; Visit #3: age 14; Visit #4: age 16) which fit the data well (CFA = .96, RMSEA = .04). Factor loadings are fixed across time, since we report standardized factor loadings, the values are slightly different across waves. For other specifications and full fit statistics, see Sup. Table 1.

**Sup. Figure 4:**
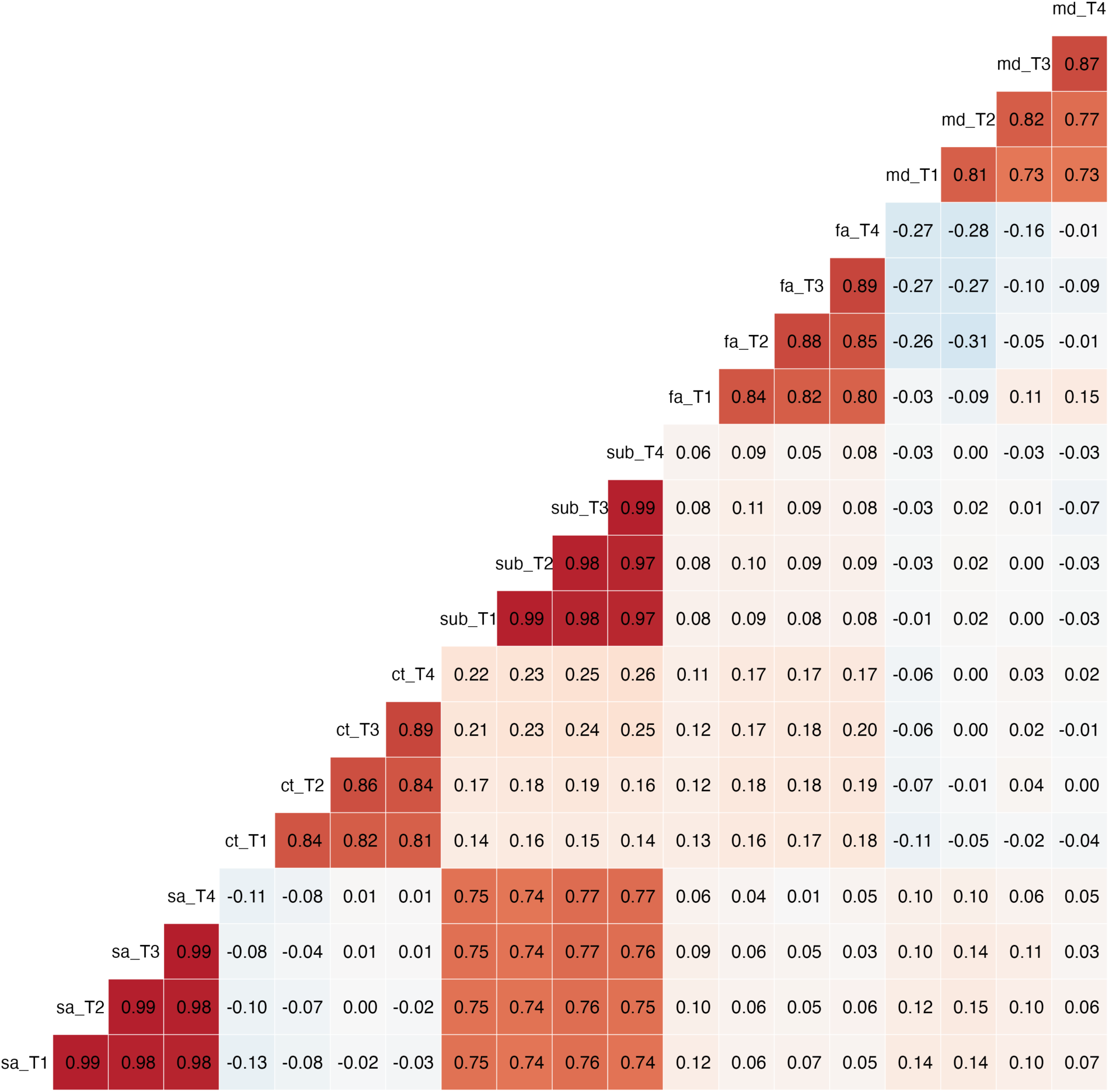
A Pearson r correlation plot of the 5 global neuroimaging measures over the 4 waves; sa = total surface area, ct = mean cortical thickness, sub = total subcortical volume, fa = average fractional anisotropy, md = average mean diffusivity. The four longitudinal time points are illustrated in order with the suffixes _T1, _T2, _T3, _T4.

**Sup. Figure 5:**
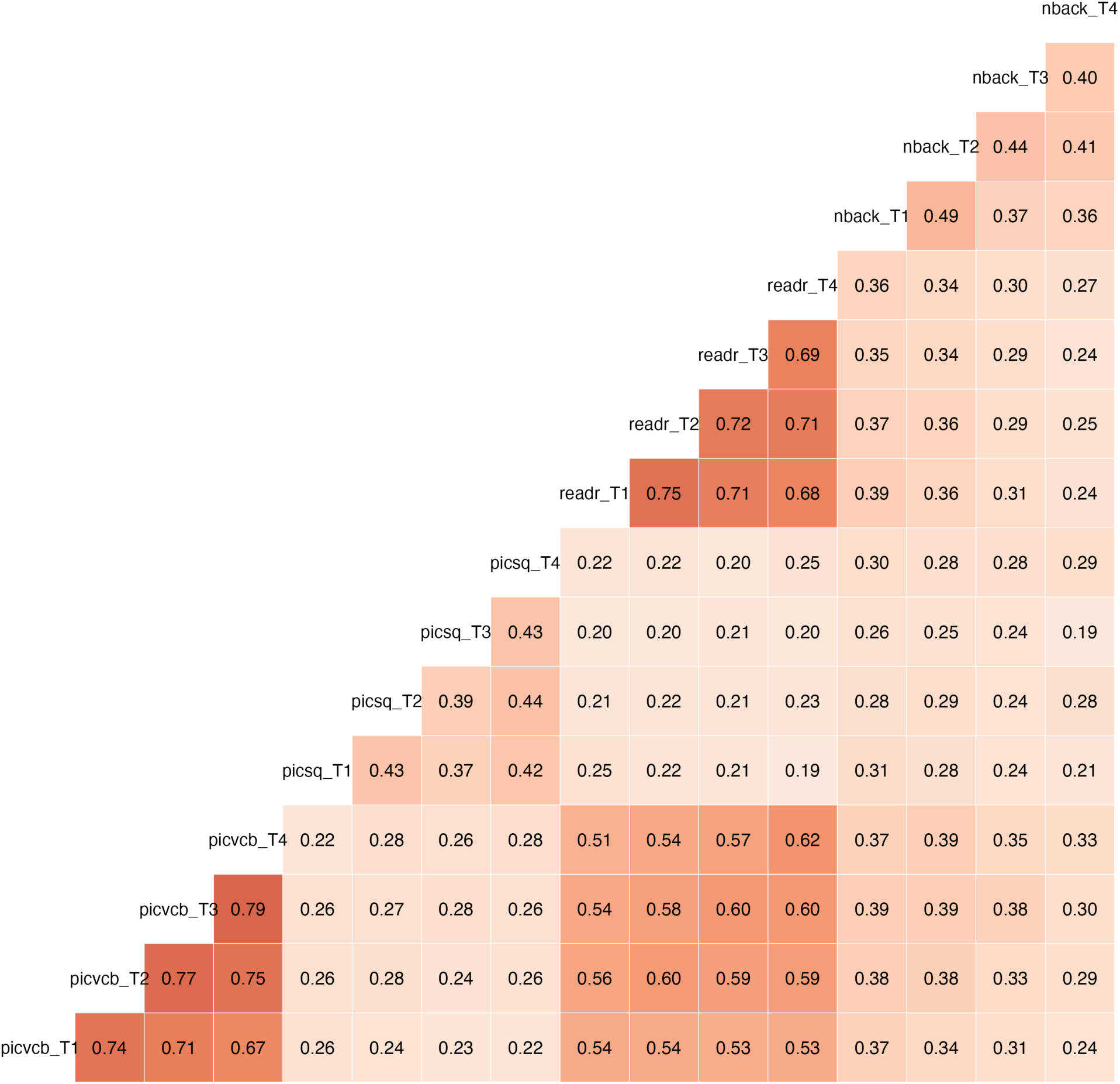
A Pearson r correlation plot of the 4 cognitive tasks over the 4 waves used to fit a strict measurement invariant CFA (see Sup. Figure 2). Task abbreviations are as follows: picvcb = Picture Vocabulary; Picsq = Picture Sequence Memory; readr = Oral Reading Recognition; nback = Working Memory nback. The four longitudinal time points are illustrated in order with the suffixes _T1, _T2, _T3, _T4.

**Sup. Figure 6:**
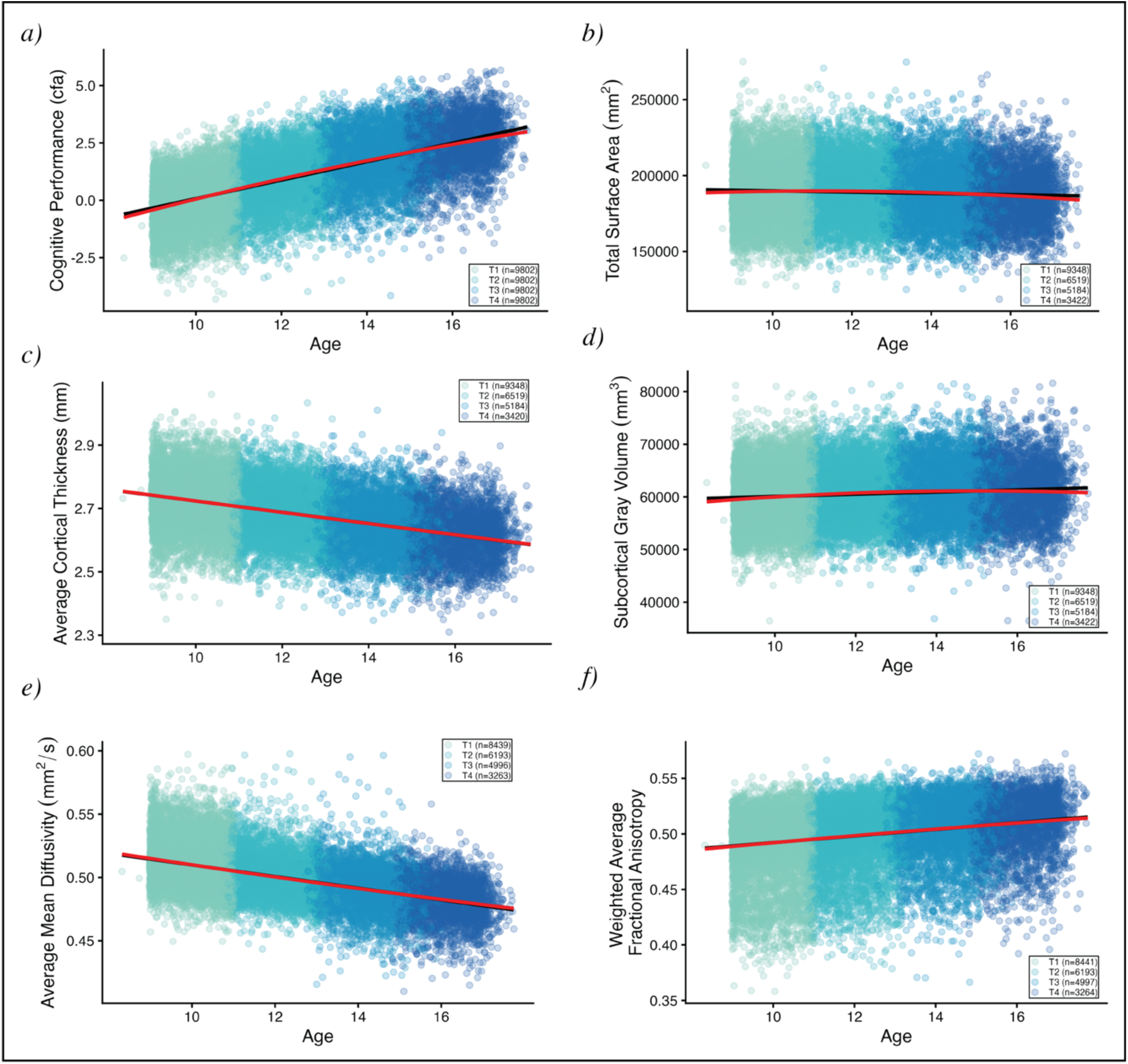
Linear (black line) and quadratic (red line) fits on cognitive performance and our five global neuroimaging measures. The addition of quadratic terms as an a priori robustness analysis (Equation 4) meaningfully increased model fit (via improved ΔAIC & ΔBIC) when compared to a linear parameterization (Equation 3) for all variables except Cortical Thickness. Nevertheless, within this age range (10 to 16 years), average trajectories are generally quite linear, as seen by the lack of major divergence between the linear (black) and quadratic (red) lines. See Sup. Figure 8 for the coefficient estimates of these trajectories per ROI.

**Sup. Figure 7:**
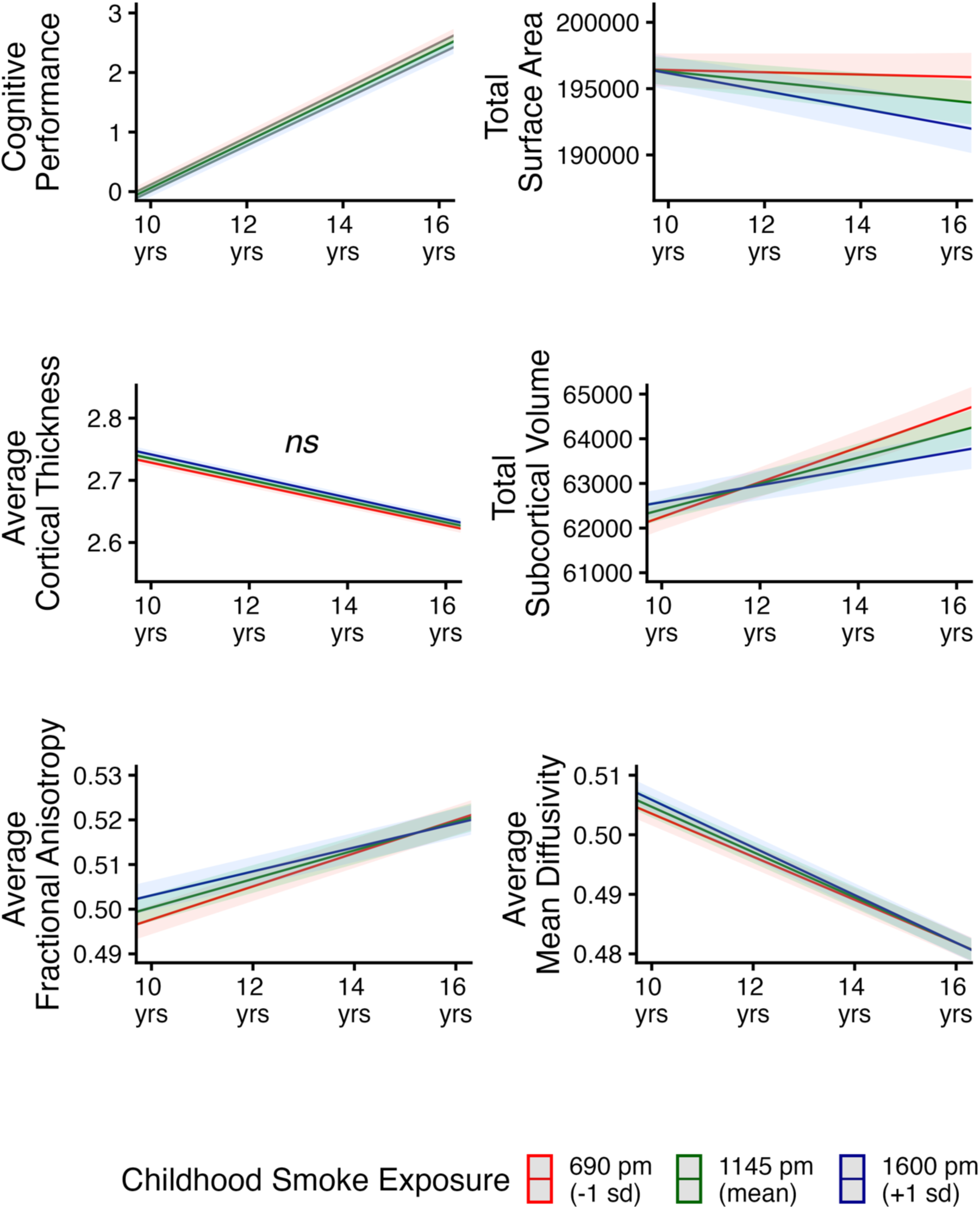
Interaction plots illustrating how childhood smoke exposure affects cognitive performance and our five global neuroimaging measures over adolescence (4 timepoints coded as age). Three CSE example trajectories are plotted using 1SD below the mean (red), mean exposure (green), and 1SD above the mean amount of exposure (blue).

**Sup. Figure 8:**
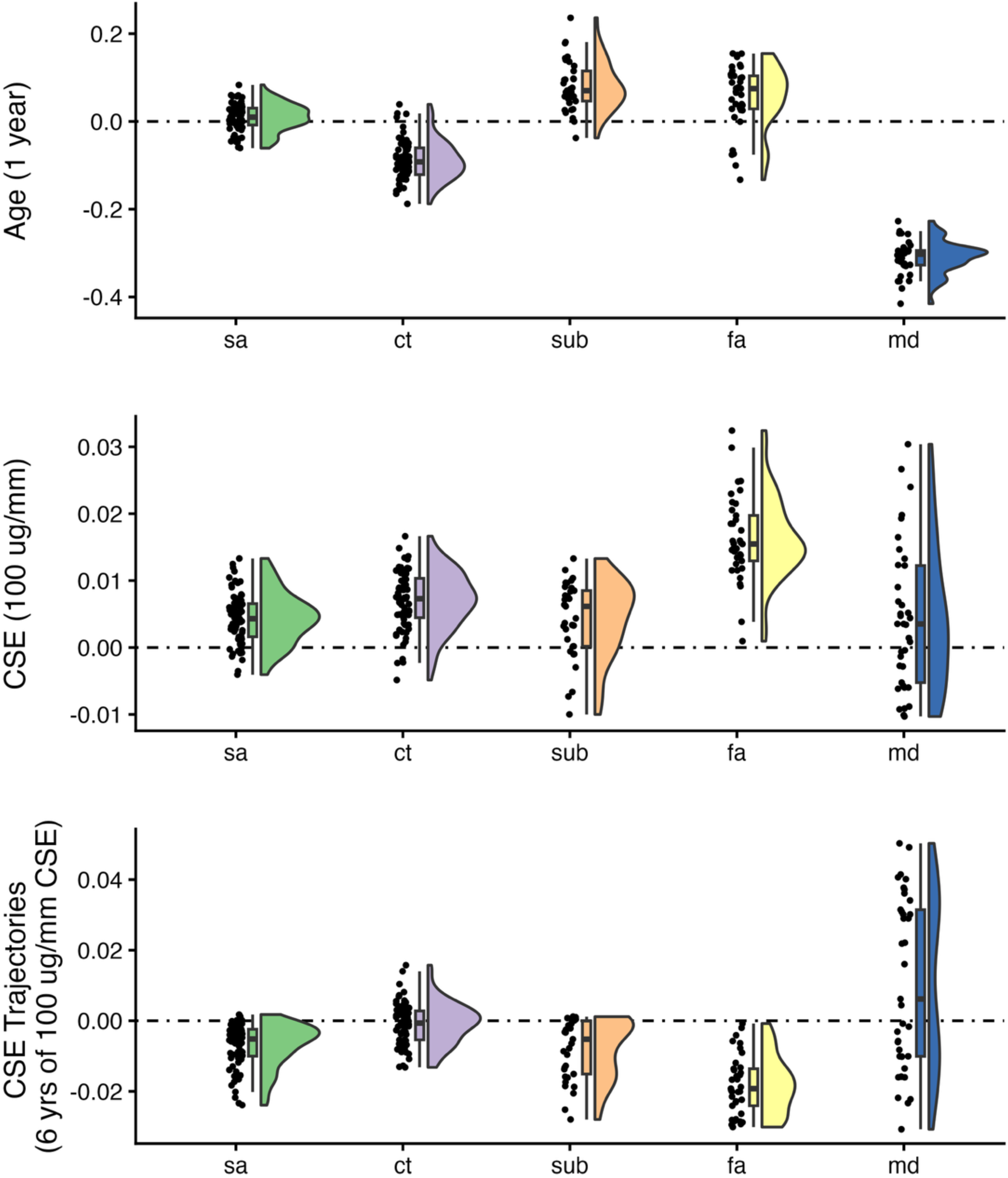
Region of interest coefficients from our main longitudinal mixed effects model (Equation 1 in methods) with childhood smoke exposure (CSE) across five structural neural modalities [sa=Surface Area, ct=Cortical Thickness, sub=Subcortical Volume, fa = fractional anisotropy, md = mean diffusivity]. For plotting purposes, the neural modalities were standardized per ROI (mean =0, SD =1). Age illustrates the coefficient of one year of development across adolescence on each ROI, while CSE illustrates the effect of CSE (in units of 100 ug/mm) on children at baseline (cross-sectionally; 10 years of age). Lastly, CSE-trajectories is the interaction of the two terms showing how additional CSE exposure (in units of 100 ug/mm) impacts the development of each ROI over the course of adolescence (6 years). Note that some ROIs were fit without a random slope for age (see Sup. Tables 10 – 14 for effect sizes in raw units, model specification, and FDR corrected significance).

**Sup Table 1.**
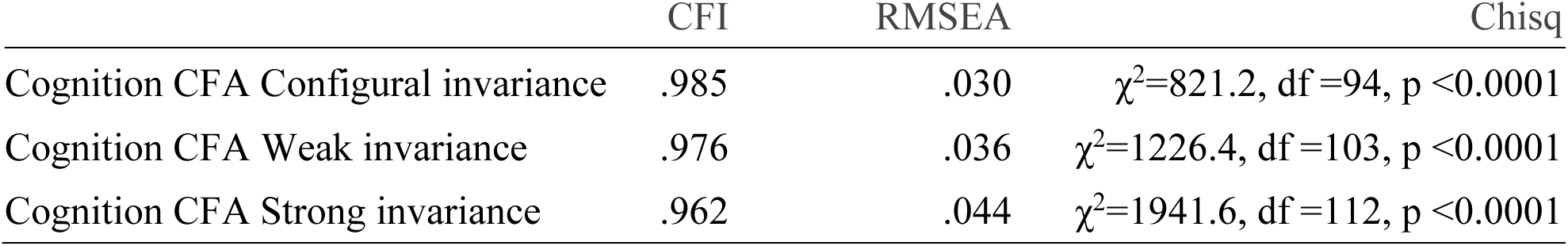
Caption: Longitudinal confirmatory factor analysis (CFA) model fit for the four cognitive tasks across four timepoints. Weak invariance indicates that the loading of tasks are fixed across time, whereas strong invariance includes additionally intercepts to be fixed across time. Factor scores from the strong invariance CFA are used for primary inference.

**Sup Table 2.** Caption: We compare two linear mixed effects models, both of which have random linear effects for age (coding for time in this design) and random intercept for participant nested within site. For subjects with at least 3 timepoints, we compared a baseline model with only a linear term for age to a model that included an additional fixed orthogonal mean-centered quadratic term. This was done to test for the presence of non-linearities in our global measures before conducting robustness analysis, Equation 4. For global FA and MD, random linear slopes did not converge for the main model (Equation 1 in Methods), we therefore report results without them. Only CT and FA did not show an improved model fit with the addition of a quadratic term. Nevertheless, even for the modalities that demonstrated improved quadratic fits, examining Figure 6 illustrates their trends as predominantly linear in this age range of adolescence (indicated by the substantial overlap between the linear black and quadratic red line).

| Y (Outcome) | $\Delta AIC$ | $\Delta BIC$ | Chisq |
| --- | --- | --- | --- |
| Cognitive Performance | 8225 | 8216 | $\chi^2=8226.7$, $\Delta df=1$, $p < 0.0001$ |
| Total Surface Area | 1885 | 1877 | $\chi^2=1887.1$, $\Delta df=1$, $p < 0.0001$ |
| Mean Cortical Thickness | -2 | -10 | $\chi^2=.002$, $\Delta df=1$, $p > 0.05$ |
| Total Subcortical Volume | 1970 | 1962 | $\chi^2=1971.7$, $\Delta df=1$, $p < 0.0001$ |
| Average Fractional Anisotropy | 1 | -7 | $\chi^2=2.8$, $\Delta df=1$, $p > 0.05$ |
| Average Mean Diffusivity | 51 | 44 | $\chi^2=52.9$, $\Delta df=1$, $p < 0.0001$ |

**Sup Table 3.** Caption: Cognitive performance factor scores (standardized by the first time point) are fit in a longitudinal mixed effects model with a random slope for time (i.e., age) and random intercepts for site and subject. In all models, age is set to the average age of the first time point (10 years old). The p-values of main effects with interactions change when centered; we report p-values on the raw beta effects (i.e., non-standardized).

| Predictors | Cognitive Performance |  |  |
| --- | --- | --- | --- |
| | Estimates ($\beta$) | std. Error | p |
| (Intercept) | 0.06564 (-0.16) | 0.08087 | 0.417 |
| Time (Age 1yr) | 0.39339 (0.63) | 0.00621 | <0.001 |
| PSE PM<sub>2.5</sub> (10 $\mu\text{g}/\text{m}^3$) | -0.00258 (-0.02) | 0.00105 | 0.014 |
| SES | 0.55626 (0.43) | 0.00996 | <0.001 |
| Mean Res PM<sub>2.5</sub> | 0.00066 (0.00) | 0.00932 | 0.944 |
| Time × PSE PM<sub>2.5</sub> (10 $\mu\text{g}/\text{m}^3$) | -0.00014 (-0.00) | 0.00008 | 0.088 |
| Time × SES | 0.01463 (0.02) | 0.00079 | <0.001 |
| Time × Mean Res PM<sub>2.5</sub> | 0.00019 (0.00) | 0.00072 | 0.790 |
| Random Effects |  |  |  |
| $\sigma^2$ | 0.02 | | |
| $\tau_{00}$ participant_id:site | 0.66 | | |
| $\tau_{00}$ site | 0.02 | | |
| $\tau_{11}$ participant_id:site.Time | 0.22 | | |
| $\tau_{11}$ site.Time | 0.01 | | |
| $\rho_{01}$ participant_id:site | 0.40 | | |
| $\rho_{01}$ site | 0.14 | | |
| ICC | 0.98 |  |  |
| $N_{\text{participant\_id}}$ | 8464 | | |
| $N_{\text{site}}$ | 22 | | |
| Observations | 26685 |  |  |
| Marginal $R^2$ / Conditional $R^2$ | 0.582 / 0.991 | | |
| AIC | 15307.796 |  |  |

**Supplementary Table 4:** CSE Cognitive Performance Results.

| Predictors | Cognitive Performance |  |  |
| --- | --- | --- | --- |
| | Estimates ($\beta$) | std. Error | p |
| (Intercept) | 0.17729 (-0.16) | 0.10173 | 0.08290 |
| Time (Age 1yr) | 0.40400 (0.63) | 0.00765 | <0.001 |
| CSE PM<sub>2.5</sub> (10 $\mu\text{g}/\text{m}^3$) | -0.00120 (-0.05) | 0.00052 | 0.023 |
| SES | 0.54804 (0.42) | 0.00957 | <0.001 |
| Res_PM<sub>2.5</sub> | -0.00026 (0.00) | 0.00904 | 0.97747 |
| Time × CSE PM<sub>2.5</sub> (10 $\mu\text{g}/\text{m}^3$) | -0.00015 (-0.01) | 0.00004 | <0.001 |
| Time × SES | 0.01377 (0.02) | 0.00076 | <0.001 |
| Time × Mean Res PM<sub>2.5</sub> | 0.00037 (0.00) | 0.00070 | 0.59728 |
| Random Effects |  |  |  |
| $\sigma^2$ | 0.02 | | |
| $\tau_{00}$ participant_id:site | 0.66 | | |
| $\tau_{00}$ site | 0.03 | | |
| $\tau_{11}$ participant_id:site.Time | 0.22 | | |
| $\tau_{11}$ site.Time | 0.02 | | |
| $\rho_{01}$ participant_id:site | 0.39 | | |
| $\rho_{01}$ site | 0.35 | | |
| ICC | 0.98 |  |  |
| N participant_id | 9372 |  |  |
| N site | 22 |  |  |
| Observations | 29919 |  |  |
| Marginal R<sup>2</sup> / Conditional R<sup>2</sup> | 0.581 / 0.991 |  |  |
| AIC | 17316.162 |  |  |

**Supplementary Table 5:**
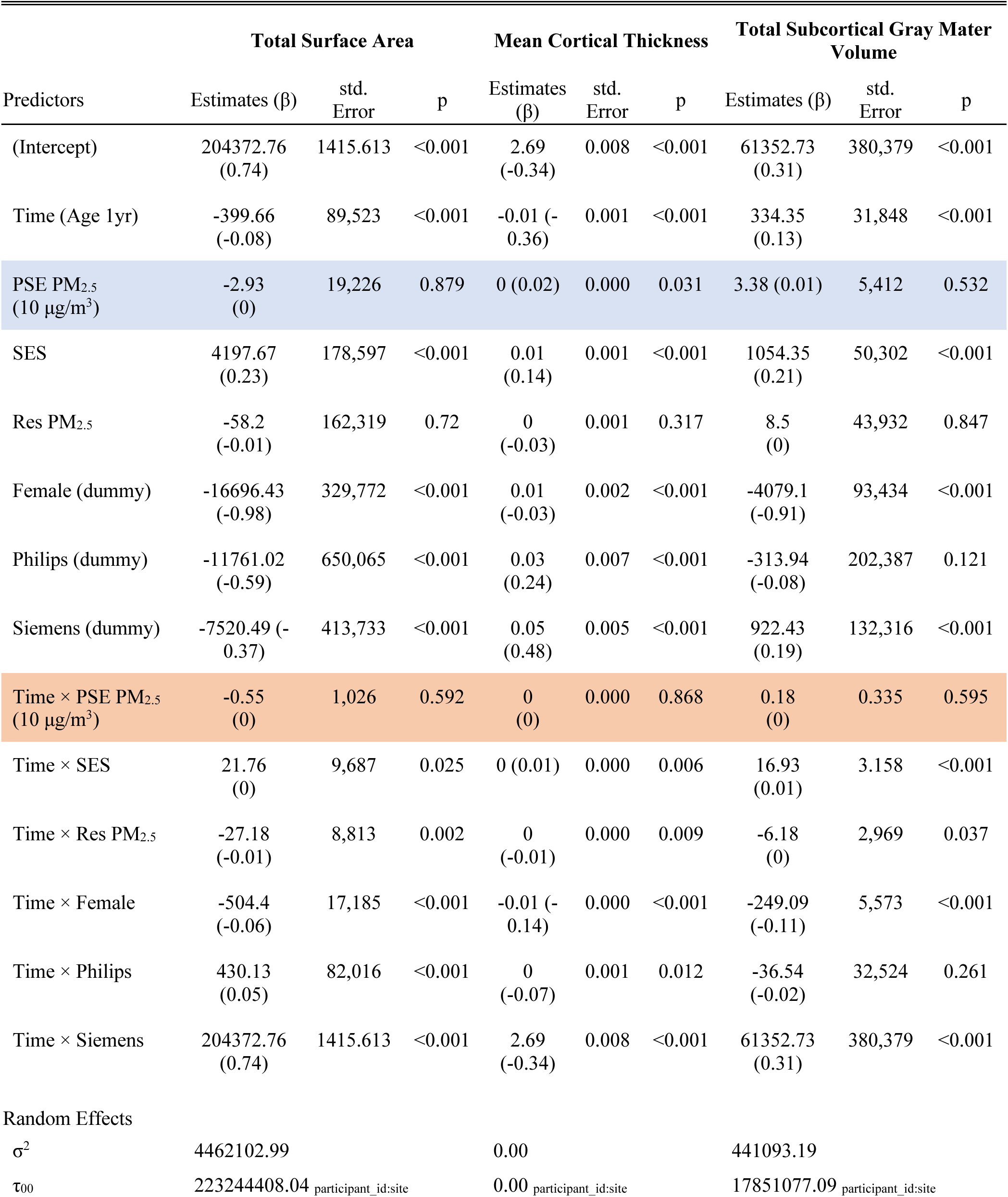
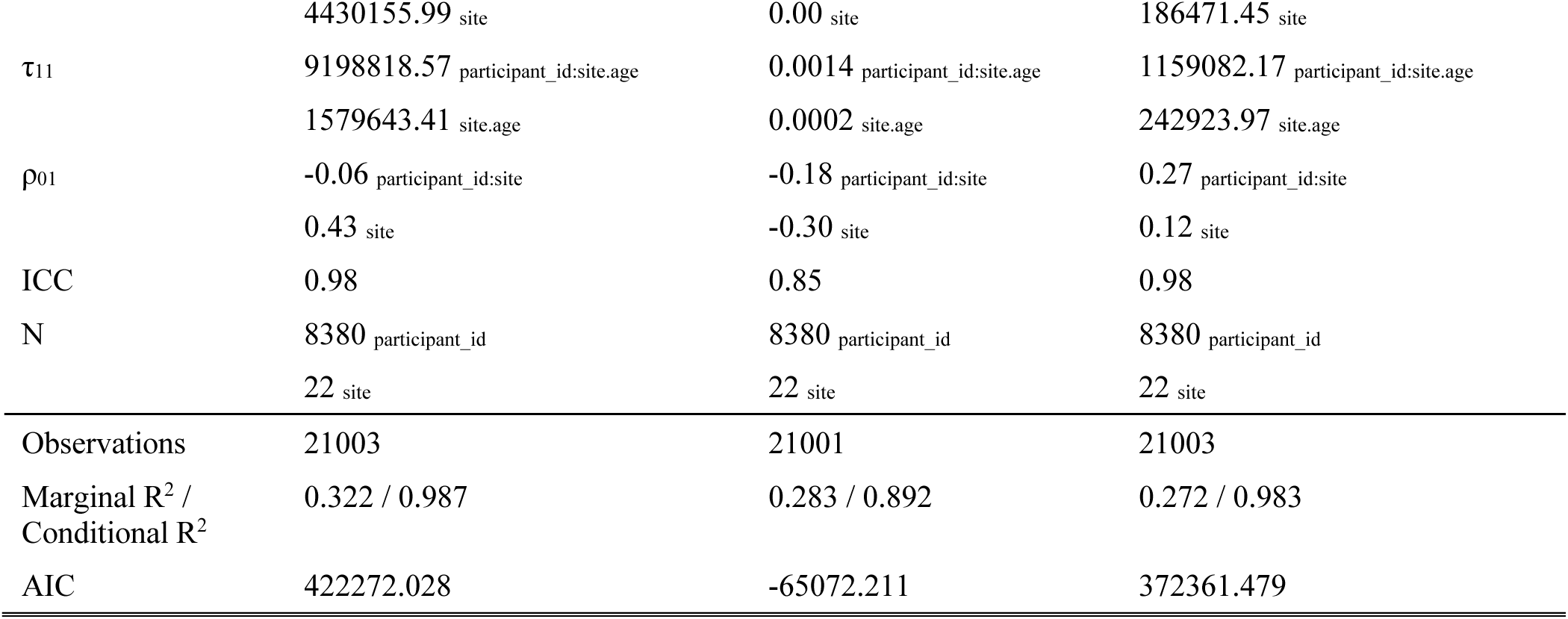
PSE Global Gray Matter Results.

**Supplementary Table 6:**
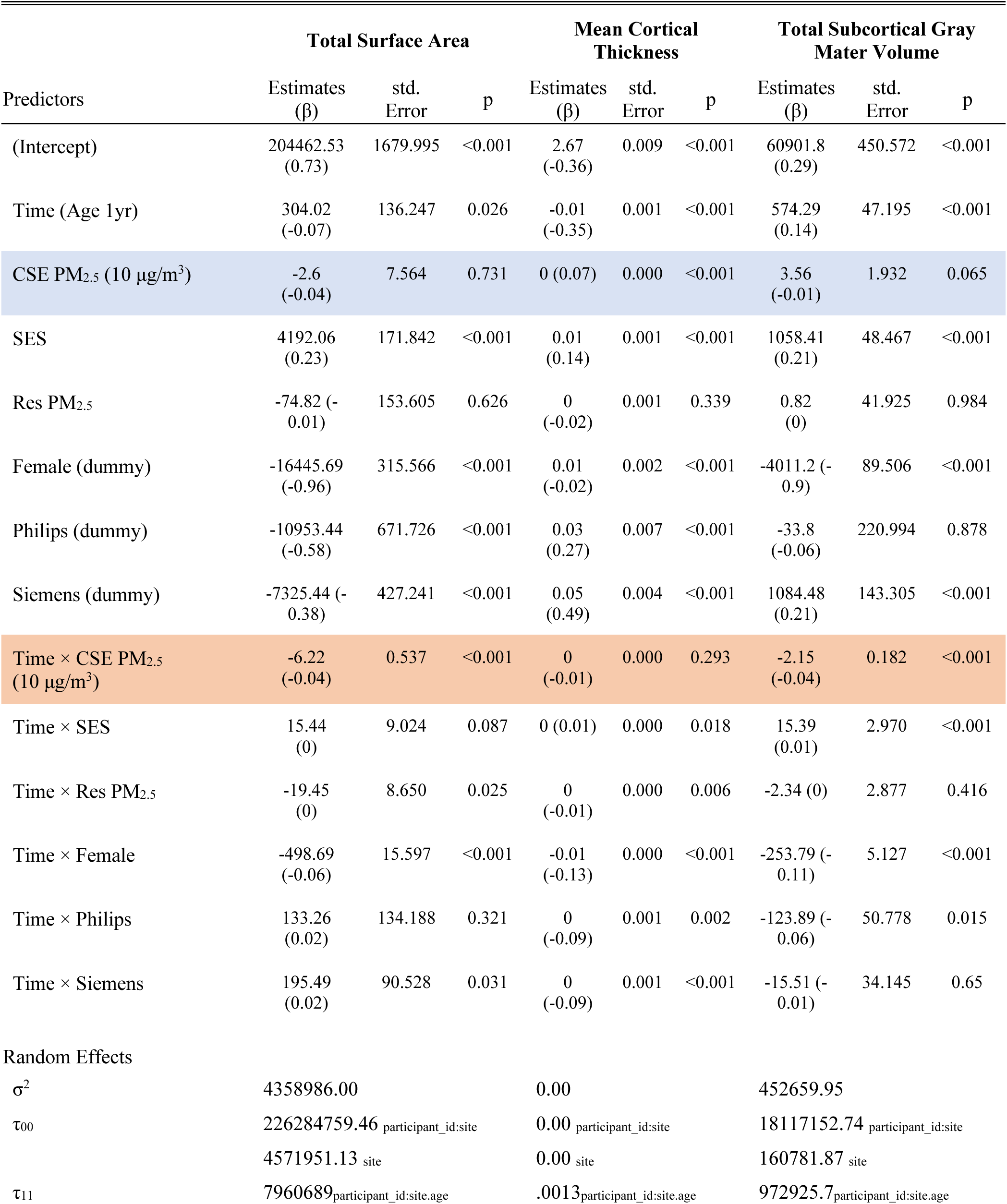
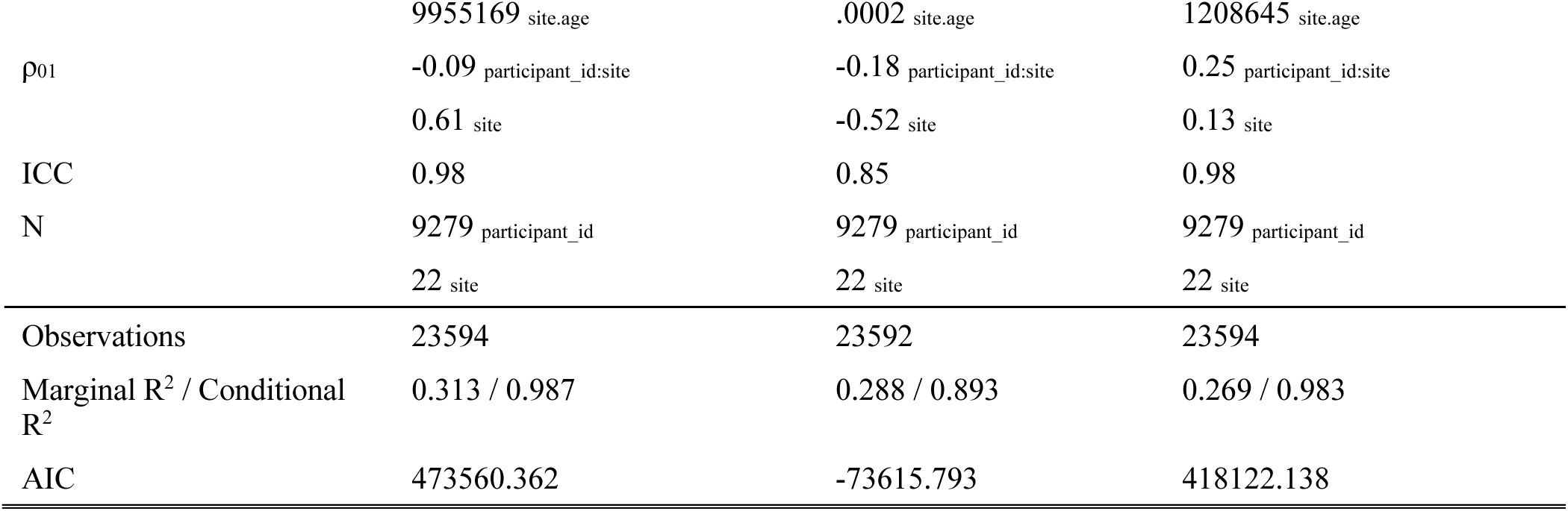
CSE Global Gray Matter Results.

| Predictors | Total Surface Area |  |  | Mean Cortical Thickness |  |  | Total Subcortical Gray Mater Volume |  |  |
| --- | --- | --- | --- | --- | --- | --- | --- | --- | --- |
|  | Estimates (β) | std. Error | p | Estimates (β) | std. Error | p | Estimates (β) | std. Error | p |
| (Intercept) | 204462.53 (0.73) | 1679.995 | <0.001 | 2.67 (-0.36) | 0.009 | <0.001 | 60901.8 (0.29) | 450.572 | <0.001 |
| Time (Age 1yr) | 304.02 (-0.07) | 136.247 | 0.026 | -0.01 (-0.35) | 0.001 | <0.001 | 574.29 (0.14) | 47.195 | <0.001 |
| CSE PM<sub>2.5</sub> (10 µg/m<sup>3</sup>) | -2.6 (-0.04) | 7.564 | 0.731 | 0 (0.07) | 0.000 | <0.001 | 3.56 (-0.01) | 1.932 | 0.065 |
| SES | 4192.06 (0.23) | 171.842 | <0.001 | 0.01 (0.14) | 0.001 | <0.001 | 1058.41 (0.21) | 48.467 | <0.001 |
| Res PM<sub>2.5</sub> | -74.82 (-0.01) | 153.605 | 0.626 | 0 (-0.02) | 0.001 | 0.339 | 0.82 (0) | 41.925 | 0.984 |
| Female (dummy) | -16445.69 (-0.96) | 315.566 | <0.001 | 0.01 (-0.02) | 0.002 | <0.001 | -4011.2 (-0.9) | 89.506 | <0.001 |
| Philips (dummy) | -10953.44 (-0.58) | 671.726 | <0.001 | 0.03 (0.27) | 0.007 | <0.001 | -33.8 (-0.06) | 220.994 | 0.878 |
| Siemens (dummy) | -7325.44 (-0.38) | 427.241 | <0.001 | 0.05 (0.49) | 0.004 | <0.001 | 1084.48 (0.21) | 143.305 | <0.001 |
| Time × CSE PM<sub>2.5</sub> (10 µg/m<sup>3</sup>) | -6.22 (-0.04) | 0.537 | <0.001 | 0 (-0.01) | 0.000 | 0.293 | -2.15 (-0.04) | 0.182 | <0.001 |
| Time × SES | 15.44 (0) | 9.024 | 0.087 | 0 (0.01) | 0.000 | 0.018 | 15.39 (0.01) | 2.970 | <0.001 |
| Time × Res PM<sub>2.5</sub> | -19.45 (0) | 8.650 | 0.025 | 0 (-0.01) | 0.000 | 0.006 | -2.34 (0) | 2.877 | 0.416 |
| Time × Female | -498.69 (-0.06) | 15.597 | <0.001 | -0.01 (-0.13) | 0.000 | <0.001 | -253.79 (-0.11) | 5.127 | <0.001 |
| Time × Philips | 133.26 (0.02) | 134.188 | 0.321 | 0 (-0.09) | 0.001 | 0.002 | -123.89 (-0.06) | 50.778 | 0.015 |
| Time × Siemens | 195.49 (0.02) | 90.528 | 0.031 | 0 (-0.09) | 0.001 | <0.001 | -15.51 (-0.01) | 34.145 | 0.65 |
| Random Effects |  |  |  |  |  |  |  |  |  |
| σ<sup>2</sup> | 4358986.00 |  |  | 0.00 |  |  | 452659.95 |  |  |
| τ<sub>00</sub> | 226284759.46<sub>participant_id:site</sub> |  |  | 0.00<sub>participant_id:site</sub> |  |  | 18117152.74<sub>participant_id:site</sub> |  |  |
|  | 4571951.13<sub>site</sub> |  |  | 0.00<sub>site</sub> |  |  | 160781.87<sub>site</sub> |  |  |
| τ<sub>11</sub> | 7960689<sub>participant_id:site:age</sub> |  |  | .0013<sub>participant_id:site:age</sub> |  |  | 972925.7<sub>participant_id:site:age</sub> |  |  |

|  |  |  |  |
| --- | --- | --- | --- |
|  | 9955169 site.age | .0002 site.age | 1208645 site.age |
| $\rho_{01}$ | -0.09 participant_id:site | -0.18 participant_id:site | 0.25 participant_id:site |
|  | 0.61 site | -0.52 site | 0.13 site |
| ICC | 0.98 | 0.85 | 0.98 |
| N | 9279 participant_id | 9279 participant_id | 9279 participant_id |
|  | 22 site | 22 site | 22 site |
| Observations | 23594 | 23592 | 23594 |
| Marginal R<sup>2</sup> / Conditional R<sup>2</sup> | 0.313 / 0.987 | 0.288 / 0.893 | 0.269 / 0.983 |
| AIC | 473560.362 | -73615.793 | 418122.138 |

**Supplementary Table 7:** PSE Global White Matter Results.

| Predictors | Fractional anisotropy |  |  | Mean Diffusivity |  |  |
| --- | --- | --- | --- | --- | --- | --- |
| | Estimates ($\beta$) | std. Error | p | Estimates ($\beta$) | std. Error | p |
| (Intercept) | 0.48991 (-0.23) | 0.00251 | <0.001 | 0.53916 (1.01) | 0.00168 | <0.001 |
| Time (Age 1yr) | 0.00111 (0.06) | 0.00022 | <0.001 | -0.00721 (-0.8) | 0.00015 | <0.001 |
| PSE PM<sub>2.5</sub><br>(10 $\mu\text{g}/\text{m}^3$) | -0.00001 (0) | 0.00022 | 0.947 | -0.00021 (-0.01) | 0.00017 | 0.215 |
| SES | 0.00146 (0.05) | 0.00020 | <0.001 | 0.00035 (0.04) | 0.00015 | 0.024 |
| Res PM<sub>2.5</sub> | 0.00014 (0) | 0.00019 | 0.469 | -0.00045 (-0.03) | 0.00015 | 0.002 |
| Female (dummy) | 0.00163 (0.07) | 0.00037 | <0.001 | -0.00271 (-0.1) | 0.00028 | <0.001 |
| Philips (dummy) | -0.05875 (-1.76) | 0.00250 | <0.001 | -0.03689 (-1.9) | 0.00165 | <0.001 |
| Siemens (dummy) | 0.00958 (0.55) | 0.00163 | <0.001 | -0.03055 (-1.19) | 0.00109 | <0.001 |
| Time × PSE PM<sub>2.5</sub><br>(10 $\mu\text{g}/\text{m}^3$) | -0.00004 (0) | 0.00004 | 0.328 | 0.00002 (0) | 0.00003 | 0.355 |
| Time × SES | 0.00003 (0) | 0.00004 | 0.398 | 0.00023 (0.03) | 0.00003 | <0.001 |
| Time × Res PM<sub>2.5</sub> | -0.00004 (-0.01) | 0.00002 | 0.095 | 0.00004 (0.01) | 0.00002 | 0.034 |
| Time × Female | 0.0001 (0.01) | 0.00007 | 0.148 | 0.00031 (0.04) | 0.00005 | <0.001 |
| Time × Philips | 0.00402 (0.33) | 0.00014 | <0.001 | -0.00022 (-0.03) | 0.00009 | 0.019 |
| Time × Siemens | 0.00247 (0.2) | 0.00009 | <0.001 | 0.00307 (0.36) | 0.00006 | <0.001 |
| Random Effects |  |  |  |  |  |  |
| $\sigma^2$ | 0.00 | | | 0.00 | | |
| $\tau_{00}$ | 0.00 participant_id:site | | | 0.00 participant_id:site | | |
|  | 0.00 site |  |  | 0.00 site |  |  |
| ICC | 0.71 |  |  | 0.75 |  |  |
| N | 8171 participant_id |  |  | 8171 participant_id |  |  |
|  | 22 site |  |  | 22 site |  |  |
| Observations | 19674 |  |  | 19671 |  |  |
| Marginal $R^2$ / Conditional $R^2$ | 0.559 / 0.872 | | | 0.590 / 0.896 | | |
| AIC | -111761.882 |  |  | -124709.176 |  |  |

**Supplementary Table 8:**
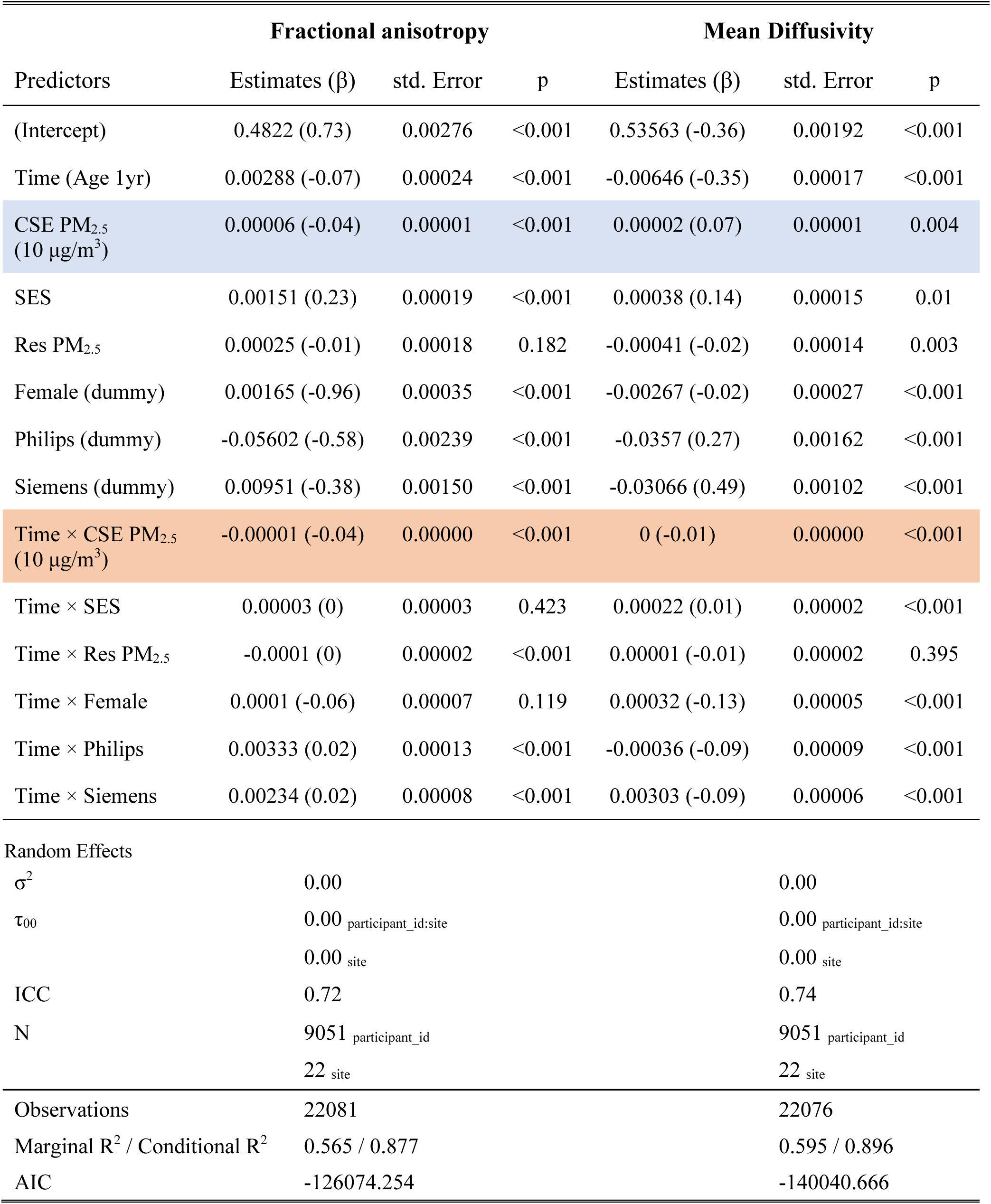
CSE Global White Matter Results.

**Sup Table 9.**
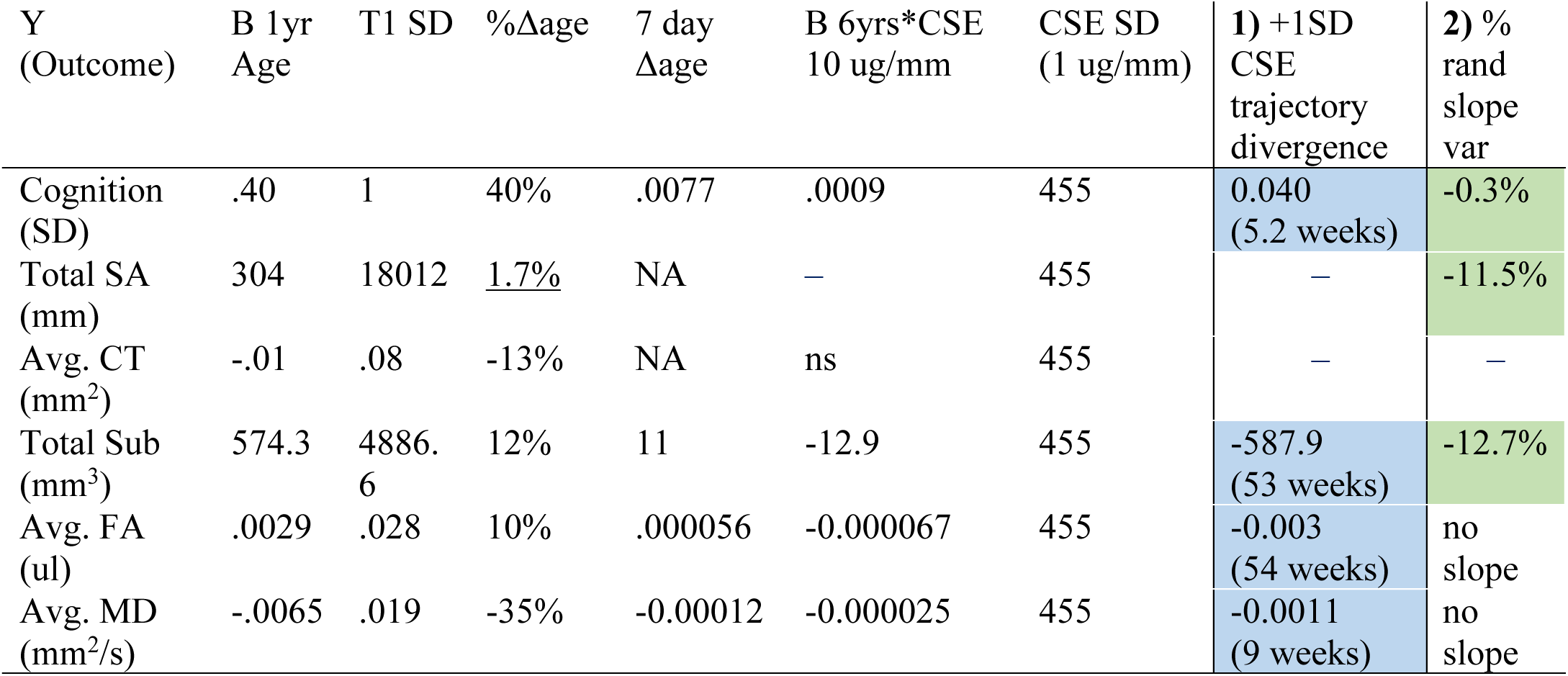
Caption: Effect size calculations of 1) weeks of divergence in blue and 2) percent of random slope variance accounted for in green; note cortical thickness is not calculated as it does not have significant trajectory effects. Weeks of divergence are calculated by taking the interaction effect size, multiplying it by 6 years (the amount of time ‘over adolescence’ in this sample), and the standard deviation of CSE. This is then divided by the weekly change (i.e., the main effect of age; 7-day Δage). This type of effect size, relies on the measure ‘changing’ – without a meaningful main effect of age, it does not make sense to talk about trajectory differences in reference to age. To get a sense of the magnitude of change, we report the percentage of change in reference to the standard deviation at T1 of that measure (%Δage). SA shows little change in our sample (see Sup. Figure 6b), only 1.7% therefore we do not calculate this measure for SA. 2) The percent of random slope variance gained is calculated by fitting the main model (Equation 1) and a model without the interaction term CSE*Age (Model 1 Null), and we then compare the amount of change in random slope variance between the two models. Multiplying this value by 100 gives us the percentage of random slope variance explained by CSE over adolescence. Models for both global FA and global MD do not converge with a random slope specification (therefore, we do not report this). Intriguingly, roughly half of their individual tracks do (see Sup. Tables 13 & 14).

**Supplementary Table 10:** Regional Surface Area Results for PSE & CSE.

| Desikan-Killiany Atlas<br>Regions (sa) | Smoke<br>Exposure | Random<br>Slopes | Model<br>Converged | Est. Main<br>effect | Est.<br>Interaction | P-val Main<br>(fdr BY) | P-val Inter<br>(fdr BY) |
| --- | --- | --- | --- | --- | --- | --- | --- |
| left caudal middle frontal | cse | TRUE | TRUE | 0.52 | **-0.07** | 0.55 | 0 |
| left fusiform | cse | TRUE | TRUE | 0.21 | **-0.1** | 1 | 0 |
| left inferior parietal | cse | TRUE | TRUE | -0.27 | **-0.11** | 1 | 0 |
| left inferior temporal | cse | TRUE | TRUE | 0.21 | **-0.15** | 1 | 0 |
| left lateral orbitofrontal | cse | TRUE | TRUE | 0.05 | **-0.13** | 1 | 0 |
| left middle temporal | cse | TRUE | TRUE | 0.13 | **-0.16** | 1 | 0 |
| left pars orbitalis | cse | TRUE | TRUE | 0.08 | **-0.02** | 1 | 0 |
| left pars opercularis | cse | TRUE | TRUE | 0.2 | **-0.05** | 1 | 0 |
| left pars triangularis | cse | TRUE | TRUE | 0.09 | **-0.07** | 1 | 0 |
| left rostral middle frontal | cse | TRUE | TRUE | 0.85 | **-0.31** | 0.91 | 0 |
| left superior frontal | cse | TRUE | TRUE | 1.03 | **-0.27** | 0.91 | 0 |
| left superior temporal | cse | TRUE | TRUE | 0.37 | **-0.15** | 1 | 0 |
| right banks of superior temporal sulcus | cse | TRUE | TRUE | 0.07 | **-0.03** | 1 | 0 |
| right caudal anterior cingulate | cse | TRUE | TRUE | 0.08 | **-0.04** | 1 | 0 |
| right fusiform | cse | TRUE | TRUE | 0.25 | **-0.09** | 1 | 0 |
| right inferior temporal | cse | TRUE | TRUE | 0.12 | **-0.14** | 1 | 0 |
| right middle temporal | cse | TRUE | TRUE | 0.03 | **-0.21** | 1 | 0 |
| right pars orbitalis | cse | TRUE | TRUE | 0.1 | **-0.02** | 1 | 0 |
| right rostral anterior cingulate | cse | TRUE | TRUE | 0.02 | **-0.02** | 1 | 0 |
| right rostral middle frontal | cse | TRUE | TRUE | 0.88 | **-0.29** | 1 | 0 |
| right superior frontal | cse | TRUE | TRUE | 1.3 | **-0.23** | 0.43 | 0 |
| right superior temporal | cse | TRUE | TRUE | 0.12 | **-0.07** | 1 | 0 |
| left banks of superior temporal sulcus | cse | TRUE | TRUE | 0 | **-0.03** | 1 | 0.01 |
| left lingual | cse | TRUE | TRUE | 0.27 | **-0.05** | 1 | 0.01 |
| left lateral occipital | cse | TRUE | TRUE | 0.38 | **-0.07** | 1 | 0.01 |
| left precentral | cse | TRUE | TRUE | 0.33 | **-0.09** | 1 | 0.01 |
| left rostral anterior cingulate | cse | TRUE | TRUE | 0.12 | **-0.03** | 1 | 0.01 |
| left supramarginal | cse | TRUE | TRUE | 0.35 | **-0.1** | 1 | 0.01 |
| right medial orbitofrontal | cse | TRUE | TRUE | 0.17 | **-0.05** | 1 | 0.01 |
| left caudal anterior cingulate | cse | TRUE | TRUE | 0.12 | **-0.01** | 0.91 | 0.02 |
| right inferior parietal | cse | TRUE | TRUE | -0.37 | **-0.09** | 1 | 0.03 |
| left insula | cse | TRUE | TRUE | 0.14 | **-0.05** | 1 | 0.04 |
| left postcentral | cse | TRUE | TRUE | 0.17 | **-0.05** | 1 | 0.04 |
| right postcentral | cse | TRUE | TRUE | 0.28 | -0.04 | 1 | 0.05 |
| right supramarginal | cse | TRUE | TRUE | 0.2 | -0.06 | 1 | 0.06 |
| right pars opercularis | cse | TRUE | TRUE | 0.19 | -0.03 | 1 | 0.08 |
| right caudal middle frontal | cse | FALSE | TRUE | 0.43 | -0.02 | 1 | 0.08 |
| right posterior cingulate | cse | TRUE | TRUE | -0.01 | -0.01 | 1 | 0.14 |
| right precentral | cse | TRUE | TRUE | 0.26 | -0.05 | 1 | 0.14 |
| right transverse temporal | cse | TRUE | TRUE | 0.03 | 0 | 1 | 0.14 |
| left frontal pole | cse | TRUE | TRUE | 0.03 | 0 | 1 | 0.21 |
| left parahippocampal | cse | TRUE | TRUE | 0.05 | -0.01 | 1 | 0.25 |
| left cuneus | cse | TRUE | TRUE | -0.02 | -0.01 | 1 | 0.26 |
| left pericalcarine | cse | TRUE | TRUE | -0.05 | -0.02 | 1 | 0.28 |
| right pars triangularis | cse | TRUE | TRUE | -0.01 | -0.02 | 1 | 0.29 |
| right lingual | cse | TRUE | TRUE | 0.28 | -0.02 | 1 | 0.3 |
| right parahippocampal | cse | TRUE | TRUE | 0.02 | -0.01 | 1 | 0.3 |
| left paracentral | cse | TRUE | TRUE | 0.02 | -0.01 | 1 | 0.6 |
| right lateral occipital | cse | TRUE | TRUE | 0.41 | -0.03 | 1 | 0.62 |
| left isthmus cingulate | cse | TRUE | TRUE | 0.09 | 0.01 | 1 | 0.74 |
| right entorhinal | cse | TRUE | TRUE | 0.08 | -0.01 | 0.55 | 0.74 |
| right paracentral | cse | TRUE | TRUE | 0.12 | -0.01 | 1 | 0.74 |
| right precuneus | cse | TRUE | TRUE | 0.26 | -0.02 | 1 | 0.79 |
| right frontal pole | cse | TRUE | TRUE | 0.05 | 0 | 1 | 0.81 |
| left entorhinal | cse | TRUE | TRUE | 0.04 | 0 | 1 | 1 |
| left medial orbitofrontal | cse | TRUE | TRUE | 0.09 | -0.01 | 1 | 1 |
| left posterior cingulate | cse | TRUE | TRUE | 0.06 | 0 | 1 | 1 |
| left precuneus | cse | TRUE | TRUE | 0.22 | 0 | 1 | 1 |
| left temporal pole | cse | TRUE | TRUE | 0.08 | 0 | 0.04 | 1 |
| left superior parietal | cse | TRUE | TRUE | 0.04 | -0.02 | 1 | 1 |
| right cuneus | cse | TRUE | TRUE | 0.01 | 0 | 1 | 1 |
| right isthmus cingulate | cse | TRUE | TRUE | -0.02 | 0 | 1 | 1 |
| right pericalcarine | cse | TRUE | TRUE | -0.02 | 0 | 1 | 1 |
| right temporal pole | cse | TRUE | TRUE | 0.09 | 0 | 0.02 | 1 |
| right superior parietal | cse | TRUE | TRUE | -0.04 | -0.02 | 1 | 1 |
| left transverse temporal | cse | FALSE | TRUE | 0.03 | 0 | 1 | 1 |
| right insula | cse | FALSE | TRUE | 0.03 | 0 | 1 | 1 |
| right lateral orbitofrontal | cse | FALSE | TRUE | -0.03 | 0 | 1 | 1 |
| left banks of superior temporal sulcus | pse | TRUE | TRUE | -0.27 | 0 | 1 | 1 |
| left caudal anterior cingulate | pse | TRUE | TRUE | 0.02 | -0.01 | 1 | 1 |
| left caudal middle frontal | pse | TRUE | TRUE | 0.54 | -0.06 | 1 | 1 |
| left cuneus | pse | TRUE | TRUE | -0.31 | 0.01 | 1 | 1 |
| left entorhinal | pse | TRUE | TRUE | 0.17 | 0.01 | 1 | 1 |
| left fusiform | pse | TRUE | TRUE | 0.21 | -0.03 | 1 | 1 |
| left isthmus cingulate | pse | TRUE | TRUE | -0.31 | 0 | 1 | 1 |
| left insula | pse | TRUE | TRUE | -0.14 | -0.08 | 1 | 1 |
| left inferior parietal | pse | TRUE | TRUE | -0.76 | 0.04 | 1 | 1 |
| left inferior temporal | pse | TRUE | TRUE | 0.13 | 0 | 1 | 1 |
| left lingual | pse | TRUE | TRUE | 0.17 | -0.02 | 1 | 1 |
| left lateral orbitofrontal | pse | TRUE | TRUE | 0.04 | 0.02 | 1 | 1 |
| left lateral occipital | pse | TRUE | TRUE | 0.28 | 0 | 1 | 1 |
| left middle temporal | pse | TRUE | TRUE | -1.19 | 0.07 | 1 | 1 |
| left paracentral | pse | TRUE | TRUE | 0.34 | -0.02 | 1 | 1 |
| left pericalcarine | pse | TRUE | TRUE | -0.14 | 0.01 | 1 | 1 |
| left posterior cingulate | pse | TRUE | TRUE | -0.16 | -0.03 | 1 | 1 |
| left frontal pole | pse | TRUE | TRUE | -0.05 | -0.01 | 1 | 1 |
| left parahippocampal | pse | TRUE | TRUE | -0.01 | 0 | 1 | 1 |
| left pars orbitalis | pse | TRUE | TRUE | -0.11 | -0.01 | 1 | 1 |
| left postcentral | pse | TRUE | TRUE | -0.13 | 0.05 | 1 | 1 |
| left pars opercularis | pse | TRUE | TRUE | 0.09 | -0.03 | 1 | 1 |
| left precuneus | pse | TRUE | TRUE | -0.05 | 0.02 | 1 | 1 |
| left precentral | pse | TRUE | TRUE | -0.2 | -0.06 | 1 | 1 |
| left pars triangularis | pse | TRUE | TRUE | -0.37 | -0.01 | 1 | 1 |
| left temporal pole | pse | TRUE | TRUE | 0.05 | -0.02 | 1 | 1 |
| left rostral anterior cingulate | pse | TRUE | TRUE | -0.06 | -0.01 | 1 | 1 |
| left rostral middle frontal | pse | TRUE | TRUE | -0.91 | -0.1 | 1 | 1 |
| left superior frontal | pse | TRUE | TRUE | 0.31 | -0.14 | 1 | 1 |
| left supramarginal | pse | TRUE | TRUE | -0.1 | 0.08 | 1 | 1 |
| left superior parietal | pse | TRUE | TRUE | -0.21 | 0.08 | 1 | 1 |
| left superior temporal | pse | TRUE | TRUE | -0.87 | 0.02 | 1 | 1 |
| left transverse temporal | pse | TRUE | TRUE | -0.05 | 0.01 | 1 | 1 |
| right banks of superior temporal sulcus | pse | TRUE | TRUE | 0.25 | -0.01 | 1 | 1 |
| right caudal anterior cingulate | pse | TRUE | TRUE | 0.07 | 0.03 | 1 | 1 |
| right caudal middle frontal | pse | TRUE | TRUE | -0.72 | -0.01 | 1 | 1 |
| right cuneus | pse | TRUE | TRUE | 0.12 | -0.01 | 1 | 1 |
| right entorhinal | pse | TRUE | TRUE | -0.01 | 0 | 1 | 1 |
| right fusiform | pse | TRUE | TRUE | 0.19 | -0.01 | 1 | 1 |
| right isthmus cingulate | pse | TRUE | TRUE | -0.09 | -0.02 | 1 | 1 |
| right insula | pse | TRUE | TRUE | -0.23 | -0.01 | 1 | 1 |
| right inferior parietal | pse | TRUE | TRUE | -0.19 | -0.15 | 1 | 1 |
| right inferior temporal | pse | TRUE | TRUE | 0.14 | 0 | 1 | 1 |
| right lingual | pse | TRUE | TRUE | -0.05 | 0 | 1 | 1 |
| right lateral orbitofrontal | pse | TRUE | TRUE | 0.08 | -0.01 | 1 | 1 |
| right medial orbitofrontal | pse | TRUE | TRUE | 0.24 | -0.03 | 1 | 1 |
| right middle temporal | pse | TRUE | TRUE | -0.35 | -0.04 | 1 | 1 |
| right paracentral | pse | TRUE | TRUE | -0.15 | 0.02 | 1 | 1 |
| right pericalcarine | pse | TRUE | TRUE | 0.08 | 0.02 | 1 | 1 |
| right posterior cingulate | pse | TRUE | TRUE | 0.05 | -0.03 | 1 | 1 |
| right parahippocampal | pse | TRUE | TRUE | 0.04 | 0 | 1 | 1 |
| right pars orbitalis | pse | TRUE | TRUE | 0 | -0.01 | 1 | 1 |
| right postcentral | pse | TRUE | TRUE | 0.24 | -0.09 | 1 | 1 |
| right pars opercularis | pse | TRUE | TRUE | 0.06 | 0.05 | 1 | 1 |
| right precuneus | pse | TRUE | TRUE | 0.61 | -0.05 | 1 | 1 |
| right precentral | pse | TRUE | TRUE | 0.04 | -0.08 | 1 | 1 |
| right pars triangularis | pse | TRUE | TRUE | 0.06 | 0.03 | 1 | 1 |
| right temporal pole | pse | TRUE | TRUE | -0.12 | 0 | 1 | 1 |
| right rostral anterior cingulate | pse | TRUE | TRUE | 0.01 | -0.01 | 1 | 1 |
| right rostral middle frontal | pse | TRUE | TRUE | -1.37 | 0.12 | 1 | 1 |
| right superior frontal | pse | TRUE | TRUE | -0.2 | 0.04 | 1 | 1 |
| right supramarginal | pse | TRUE | TRUE | -0.75 | -0.08 | 1 | 1 |
| right superior parietal | pse | TRUE | TRUE | 0.1 | -0.06 | 1 | 1 |
| right superior temporal | pse | TRUE | TRUE | -0.6 | -0.06 | 1 | 1 |
| right transverse temporal | pse | TRUE | TRUE | -0.06 | 0 | 1 | 1 |
| left medial orbitofrontal | pse | FALSE | TRUE | 0.02 | 0.01 | 1 | 1 |
| right lateral occipital | pse | FALSE | TRUE | 0.17 | -0.06 | 1 | 1 |
| right frontal pole | pse | FALSE | TRUE | 0.05 | 0 | 1 | 1 |

**Supplementary Table 11:** Regional Cortical Thickness Results for PSE & CSE :

| Desikan-Killiany Atlas<br>Regions (sa) | Smoke<br>Exposure | Random<br>Slopes | Model<br>Converged | Est. Main<br>effect | Est.<br>Interaction | P-val Main<br>(fdr BY) | P-val Inter<br>(fdr BY) |
| --- | --- | --- | --- | --- | --- | --- | --- |
| left posterior cingulate | cse | FALSE | TRUE | 0 | **0** | 1 | 0 |
| left superior parietal | cse | FALSE | TRUE | 0 | **0** | 0.81 | 0 |
| left medial orbitofrontal | cse | FALSE | TRUE | 0 | **0** | 1 | 0.01 |
| left paracentral | cse | TRUE | TRUE | 0 | 0 | 1 | 0.08 |
| right posterior cingulate | cse | TRUE | TRUE | 0 | 0 | 0.22 | 0.09 |
| right insula | cse | FALSE | TRUE | 0 | 0 | 1 | 0.13 |
| left cuneus | cse | TRUE | TRUE | 0 | 0 | 1 | 0.18 |
| right isthmus cingulate | cse | TRUE | TRUE | 0 | 0 | 0.22 | 0.18 |
| left isthmus cingulate | cse | TRUE | TRUE | 0 | 0 | 0.08 | 0.28 |
| left lingual | cse | TRUE | TRUE | 0 | 0 | 1 | 0.28 |
| left precentral | cse | TRUE | TRUE | 0 | 0 | 1 | 0.39 |
| left rostral anterior cingulate | cse | FALSE | FALSE | 0 | 0 | 1 | 0.52 |
| right lingual | cse | TRUE | TRUE | 0 | 0 | 1 | 0.56 |
| right paracentral | cse | TRUE | TRUE | 0 | 0 | 1 | 0.56 |
| left inferior parietal | cse | TRUE | TRUE | 0 | 0 | 0.91 | 0.83 |
| right middle temporal | cse | TRUE | TRUE | 0 | 0 | 0.54 | 0.98 |
| left caudal anterior cingulate | cse | TRUE | TRUE | 0 | 0 | 1 | 1 |
| left caudal middle frontal | cse | TRUE | TRUE | 0 | 0 | 0.51 | 1 |
| left entorhinal | cse | TRUE | TRUE | 0 | 0 | 1 | 1 |
| left fusiform | cse | TRUE | TRUE | 0 | 0 | 1 | 1 |
| left inferior temporal | cse | TRUE | TRUE | 0 | 0 | 1 | 1 |
| left lateral orbitofrontal | cse | TRUE | TRUE | 0 | 0 | 0.51 | 1 |
| left lateral occipital | cse | TRUE | TRUE | 0 | 0 | 1 | 1 |
| left pericalcarine | cse | TRUE | TRUE | 0 | 0 | 1 | 1 |
| left frontal pole | cse | TRUE | TRUE | 0 | 0 | 0.25 | 1 |
| left parahippocampal | cse | TRUE | TRUE | 0 | 0 | 0.81 | 1 |
| left postcentral | cse | TRUE | TRUE | 0 | 0 | 1 | 1 |
| left pars opercularis | cse | TRUE | TRUE | 0 | 0 | 0.81 | 1 |
| left precuneus | cse | TRUE | TRUE | 0 | 0 | 1 | 1 |
| left pars triangularis | cse | TRUE | TRUE | 0 | 0 | 1 | 1 |
| left temporal pole | cse | TRUE | TRUE | 0 | 0 | 1 | 1 |
| left rostral middle frontal | cse | TRUE | TRUE | 0 | 0 | 1 | 1 |
| left supramarginal | cse | TRUE | TRUE | 0 | 0 | 1 | 1 |
| left superior temporal | cse | TRUE | TRUE | 0 | 0 | 0.37 | 1 |
| left transverse temporal | cse | TRUE | TRUE | 0 | 0 | 0.22 | 1 |
| right banks of superior temporal sulcus | cse | TRUE | TRUE | 0 | 0 | 1 | 1 |
| right caudal anterior cingulate | cse | TRUE | TRUE | 0 | 0 | 1 | 1 |
| right caudal middle frontal | cse | TRUE | TRUE | 0 | 0 | 0.79 | 1 |
| right cuneus | cse | TRUE | TRUE | 0 | 0 | 1 | 1 |
| right entorhinal | cse | TRUE | TRUE | 0 | 0 | 1 | 1 |
| right fusiform | cse | TRUE | TRUE | 0 | 0 | 1 | 1 |
| right lateral orbitofrontal | cse | TRUE | TRUE | 0 | 0 | 0.76 | 1 |
| right pericalcarine | cse | TRUE | TRUE | 0 | 0 | 1 | 1 |
| right frontal pole | cse | TRUE | TRUE | 0 | 0 | 1 | 1 |
| right parahippocampal | cse | TRUE | TRUE | 0 | 0 | 0.3 | 1 |
| right pars orbitalis | cse | TRUE | TRUE | 0 | 0 | 1 | 1 |
| right postcentral | cse | TRUE | TRUE | 0 | 0 | 1 | 1 |
| right pars opercularis | cse | TRUE | TRUE | 0 | 0 | 1 | 1 |
| right precuneus | cse | TRUE | TRUE | 0 | 0 | 0.3 | 1 |
| right precentral | cse | TRUE | TRUE | 0 | 0 | 1 | 1 |
| right pars triangularis | cse | TRUE | TRUE | 0 | 0 | 1 | 1 |
| right temporal pole | cse | TRUE | TRUE | 0 | 0 | 1 | 1 |
| right rostral anterior cingulate | cse | TRUE | TRUE | 0 | 0 | 1 | 1 |
| right rostral middle frontal | cse | TRUE | TRUE | 0 | 0 | 1 | 1 |
| right superior frontal | cse | TRUE | TRUE | 0 | 0 | 0.81 | 1 |
| right supramarginal | cse | TRUE | TRUE | 0 | 0 | 1 | 1 |
| right superior parietal | cse | TRUE | TRUE | 0 | 0 | 1 | 1 |
| right superior temporal | cse | TRUE | TRUE | 0 | 0 | 0.3 | 1 |
| right transverse temporal | cse | TRUE | TRUE | 0 | 0 | 1 | 1 |
| left banks of superior temporal sulcus | cse | FALSE | TRUE | 0 | 0 | 1 | 1 |
| left insula | cse | FALSE | TRUE | 0 | 0 | 1 | 1 |
| left middle temporal | cse | FALSE | TRUE | 0 | 0 | 1 | 1 |
| left pars orbitalis | cse | FALSE | TRUE | 0 | 0 | 0.25 | 1 |
| left superior frontal | cse | FALSE | TRUE | 0 | 0 | 0.54 | 1 |
| right inferior parietal | cse | FALSE | TRUE | 0 | 0 | 0.25 | 1 |
| right inferior temporal | cse | FALSE | TRUE | 0 | 0 | 0.25 | 1 |
| right lateral occipital | cse | FALSE | TRUE | 0 | 0 | 0.26 | 1 |
| right medial orbitofrontal | cse | FALSE | TRUE | 0 | 0 | 0.81 | 1 |
| left banks of superior temporal sulcus | pse | TRUE | TRUE | 0 | -0.03 | 0.07 | 1 |
| left caudal anterior cingulate | pse | TRUE | TRUE | 0 | 0.06 | 1 | 1 |
| left caudal middle frontal | pse | TRUE | TRUE | 0 | 0.01 | 1 | 1 |
| left cuneus | pse | TRUE | TRUE | 0 | 0.01 | 1 | 1 |
| left entorhinal | pse | TRUE | TRUE | 0 | -0.05 | 1 | 1 |
| left fusiform | pse | TRUE | TRUE | 0 | 0 | 1 | 1 |
| left inferior parietal | pse | TRUE | TRUE | 0 | 0 | 1 | 1 |
| left inferior temporal | pse | TRUE | TRUE | 0 | 0 | 0.42 | 1 |
| left lingual | pse | TRUE | TRUE | 0 | 0.03 | 1 | 1 |
| left lateral orbitofrontal | pse | TRUE | TRUE | 0 | -0.02 | 1 | 1 |
| left lateral occipital | pse | TRUE | TRUE | 0 | 0.01 | 1 | 1 |
| left medial orbitofrontal | pse | TRUE | TRUE | 0 | 0.03 | 1 | 1 |
| left paracentral | pse | TRUE | TRUE | 0 | -0.01 | 0.91 | 1 |
| left pericalcarine | pse | TRUE | TRUE | 0 | 0.01 | 1 | 1 |
| left frontal pole | pse | TRUE | TRUE | 0 | 0 | 1 | 1 |
| left parahippocampal | pse | TRUE | TRUE | 0 | -0.01 | 1 | 1 |
| left pars opercularis | pse | TRUE | TRUE | 0 | 0 | 0.45 | 1 |
| left precuneus | pse | TRUE | TRUE | 0 | 0 | 0.91 | 1 |
| left precentral | pse | TRUE | TRUE | 0 | 0.01 | 1 | 1 |
| left pars triangularis | pse | TRUE | TRUE | 0 | -0.01 | 1 | 1 |
| left temporal pole | pse | TRUE | TRUE | 0 | 0.06 | 1 | 1 |
| left rostral anterior cingulate | pse | TRUE | TRUE | 0 | 0.06 | 1 | 1 |
| left rostral middle frontal | pse | TRUE | TRUE | 0 | 0.01 | 1 | 1 |
| left superior frontal | pse | TRUE | TRUE | 0 | 0 | 1 | 1 |
| left supramarginal | pse | TRUE | TRUE | 0 | -0.02 | 0.2 | 1 |
| left superior parietal | pse | TRUE | TRUE | 0 | 0.01 | 1 | 1 |
| left superior temporal | pse | TRUE | TRUE | 0 | -0.03 | 0.07 | 1 |
| left transverse temporal | pse | TRUE | TRUE | 0 | -0.01 | 1 | 1 |
| right caudal anterior cingulate | pse | TRUE | TRUE | 0 | -0.01 | 0.91 | 1 |
| right caudal middle frontal | pse | TRUE | TRUE | 0 | -0.03 | 1 | 1 |
| right cuneus | pse | TRUE | TRUE | 0 | 0 | 1 | 1 |
| right fusiform | pse | TRUE | TRUE | 0 | 0 | 1 | 1 |
| right isthmus cingulate | pse | TRUE | TRUE | 0 | -0.01 | 1 | 1 |
| right insula | pse | TRUE | TRUE | 0 | 0.01 | 1 | 1 |
| right inferior parietal | pse | TRUE | TRUE | 0 | -0.01 | 1 | 1 |
| right inferior temporal | pse | TRUE | TRUE | 0 | -0.02 | 0.91 | 1 |
| right lingual | pse | TRUE | TRUE | 0 | 0 | 1 | 1 |
| right lateral orbitofrontal | pse | TRUE | TRUE | 0 | -0.01 | 1 | 1 |
| right lateral occipital | pse | TRUE | TRUE | 0 | 0.03 | 1 | 1 |
| right medial orbitofrontal | pse | TRUE | TRUE | 0 | 0.01 | 1 | 1 |
| right paracentral | pse | TRUE | TRUE | 0 | -0.04 | 0.07 | 1 |
| right pericalcarine | pse | TRUE | TRUE | 0 | -0.02 | 1 | 1 |
| right frontal pole | pse | TRUE | TRUE | 0 | -0.11 | 1 | 1 |
| right parahippocampal | pse | TRUE | TRUE | 0 | -0.06 | 0.91 | 1 |
| right pars orbitalis | pse | TRUE | TRUE | 0 | -0.03 | 0.91 | 1 |
| right postcentral | pse | TRUE | TRUE | 0 | 0.03 | 1 | 1 |
| right pars opercularis | pse | TRUE | TRUE | 0 | -0.01 | 1 | 1 |
| right precuneus | pse | TRUE | TRUE | 0 | -0.02 | 1 | 1 |
| right precentral | pse | TRUE | TRUE | 0 | -0.04 | 1 | 1 |
| right pars triangularis | pse | TRUE | TRUE | 0 | -0.03 | 1 | 1 |
| right rostral anterior cingulate | pse | TRUE | TRUE | 0 | 0.03 | 1 | 1 |
| right rostral middle frontal | pse | TRUE | TRUE | 0 | -0.02 | 1 | 1 |
| right superior frontal | pse | TRUE | TRUE | 0 | -0.01 | 1 | 1 |
| right supramarginal | pse | TRUE | TRUE | 0 | 0.01 | 1 | 1 |
| right superior parietal | pse | TRUE | TRUE | 0 | 0.01 | 1 | 1 |
| right superior temporal | pse | TRUE | TRUE | 0 | 0.01 | 1 | 1 |
| right transverse temporal | pse | TRUE | TRUE | 0 | 0.01 | 0.91 | 1 |
| left isthmus cingulate | pse | FALSE | TRUE | 0 | -0.02 | 0.27 | 1 |
| left insula | pse | FALSE | TRUE | 0 | 0.03 | 1 | 1 |
| left middle temporal | pse | FALSE | TRUE | 0 | -0.01 | 0.91 | 1 |
| left posterior cingulate | pse | FALSE | TRUE | 0 | -0.02 | 0.2 | 1 |
| left pars orbitalis | pse | FALSE | TRUE | 0 | -0.02 | 1 | 1 |
| left postcentral | pse | FALSE | TRUE | 0 | 0 | 1 | 1 |
| right banks of superior temporal sulcus | pse | FALSE | TRUE | 0 | -0.05 | 0.91 | 1 |
| right entorhinal | pse | FALSE | TRUE | 0 | -0.13 | 1 | 1 |
| right middle temporal | pse | FALSE | TRUE | 0 | -0.02 | 0.91 | 1 |
| right posterior cingulate | pse | FALSE | TRUE | 0 | -0.03 | 1 | 1 |
| right temporal pole | pse | FALSE | TRUE | 0 | 0.06 | 1 | 1 |

**Supplementary Table 12:** Regional Subcortical Results for PSE & CSE.

| Subcortical Region | Smoke Exposure | Random Slopes | Model Converged | Est. Main effect | P-val Main (fdr BY) | Est. Interaction | P-val Inter (fdr BY) |
| --- | --- | --- | --- | --- | --- | --- | --- |
| left cerebellum cortex | cse | TRUE | TRUE | 7.903 | 0.033 | **-2.038** | 0 |
| left cerebral white matter | cse | TRUE | TRUE | 20.321 | 0.108 | **-6.911** | 0 |
| left hippocampus | cse | TRUE | TRUE | 0.266 | 0.171 | **-0.095** | 0 |
| left ventral diencephalon | cse | TRUE | TRUE | 0.265 | 0.148 | **-0.124** | 0 |
| right cerebellum cortex | cse | TRUE | TRUE | 7.123 | 0.049 | **-1.897** | 0 |
| right cerebral white matter | cse | TRUE | TRUE | 20.064 | 0.121 | **-7.018** | 0 |
| right hippocampus | cse | TRUE | TRUE | 0.143 | 0.53 | **-0.109** | 0 |
| right putamen | cse | TRUE | TRUE | 0.354 | 0.106 | **-0.123** | 0 |
| right ventral diencephalon | cse | TRUE | TRUE | 0.381 | 0.049 | **-0.176** | 0 |
| brain stem | cse | TRUE | TRUE | 1.839 | 0.062 | **-1.021** | 0 |
| right amygdala | cse | FALSE | FALSE | 0.206 | 0.055 | -0.023 | 0 |
| left amygdala | cse | TRUE | TRUE | 0.173 | 0.179 | **-0.053** | 0.001 |
| left caudate | cse | TRUE | TRUE | 0.421 | 0.062 | **-0.051** | 0.002 |
| mid-posterior corpus callosum | cse | TRUE | TRUE | 0.073 | 0.106 | **-0.013** | 0.007 |
| right caudate | cse | TRUE | TRUE | 0.495 | 0.038 | **-0.042** | 0.011 |
| right thalamus | cse | TRUE | TRUE | 0.238 | 0.53 | **-0.107** | 0.012 |
| left putamen | cse | FALSE | TRUE | 0.212 | 0.211 | **-0.031** | 0.013 |
| posterior corpus callosum | cse | TRUE | TRUE | 0.139 | 0.033 | -0.008 | 0.083 |
| central corpus callosum | cse | TRUE | TRUE | -0.008 | 0.946 | -0.005 | 0.51 |
| mid-anterior corpus callosum | cse | TRUE | TRUE | 0.042 | 0.632 | -0.007 | 0.556 |
| left inferior lateral ventricle | cse | TRUE | TRUE | -0.14 | 0.033 | -0.004 | 0.779 |
| 4th ventricle | cse | TRUE | TRUE | -0.151 | 0.534 | 0.008 | 0.779 |
| left lateral ventricle | cse | TRUE | TRUE | 0.051 | 0.964 | 0.029 | 0.834 |
| right inferior lateral ventricle | cse | TRUE | TRUE | -0.101 | 0.049 | -0.003 | 0.834 |
| right lateral ventricle | cse | TRUE | TRUE | 0.855 | 0.375 | 0.035 | 0.834 |
| lateral ventricles | cse | TRUE | TRUE | 0.643 | 0.786 | 0.066 | 0.834 |
| anterior corpus callosum | cse | TRUE | TRUE | 0.141 | 0.088 | -0.002 | 0.839 |
| left pallidum | cse | TRUE | TRUE | 0.004 | 0.964 | 0.005 | 0.84 |
| left thalamus | cse | FALSE | FALSE | 0.38 | 0.393 | -0.005 | 0.918 |
| right accumbens area | cse | TRUE | TRUE | 0.043 | 0.25 | 0.001 | 0.953 |
| right pallidum | cse | TRUE | TRUE | -0.023 | 0.852 | 0.002 | 0.953 |
| left accumbens area | cse | TRUE | TRUE | -0.011 | 0.865 | 0 | 0.982 |
| 3rd ventricle | cse | TRUE | TRUE | -0.139 | 0.106 | 0 | 0.982 |
| left cerebellum cortex | pse | TRUE | TRUE | -1.553 | 0.952 | 1.201 | 0.272 |
| right cerebellum cortex | pse | TRUE | TRUE | -3.753 | 0.952 | 0.961 | 0.687 |
| right pallidum | pse | TRUE | TRUE | -0.047 | 0.952 | -0.085 | 0.687 |
| left amygdala | pse | TRUE | TRUE | -0.159 | 0.952 | 0.035 | 0.828 |
| left inferior lateral ventricle | pse | TRUE | TRUE | -0.028 | 0.952 | 0.036 | 0.828 |
| left lateral ventricle | pse | TRUE | TRUE | 1.045 | 0.952 | -0.182 | 0.828 |
| left pallidum | pse | TRUE | TRUE | -0.47 | 0.952 | -0.041 | 0.828 |
| right accumbens area | pse | TRUE | TRUE | 0.154 | 0.952 | 0.019 | 0.828 |
| right amygdala | pse | TRUE | TRUE | -0.125 | 0.952 | 0.029 | 0.828 |
| right hippocampus | pse | TRUE | TRUE | 0.269 | 0.952 | 0.035 | 0.828 |
| right lateral ventricle | pse | TRUE | TRUE | -0.315 | 0.952 | -0.295 | 0.828 |
| right putamen | pse | TRUE | TRUE | 0.469 | 0.952 | 0.054 | 0.828 |
| right thalamus | pse | TRUE | TRUE | 1.03 | 0.952 | -0.07 | 0.828 |
| right ventral diencephalon | pse | TRUE | TRUE | 0.118 | 0.952 | 0.06 | 0.828 |
| mid-anterior corpus callosum | pse | TRUE | TRUE | 0.011 | 0.952 | 0.016 | 0.828 |
| posterior corpus callosum | pse | TRUE | TRUE | -0.026 | 0.952 | -0.008 | 0.828 |
| mid-posterior corpus callosum | pse | TRUE | TRUE | 0.132 | 0.952 | 0.007 | 0.828 |
| lateral ventricles | pse | TRUE | TRUE | -0.427 | 0.952 | -0.456 | 0.828 |
| left hippocampus | pse | FALSE | TRUE | 0.352 | 0.952 | -0.054 | 0.828 |
| left thalamus | pse | TRUE | TRUE | 0.488 | 0.952 | -0.076 | 0.885 |
| anterior corpus callosum | pse | TRUE | TRUE | 0.022 | 0.952 | -0.008 | 0.9 |
| right cerebral white matter | pse | TRUE | TRUE | 5.859 | 0.952 | -0.712 | 0.904 |
| 3rd ventricle | pse | TRUE | TRUE | -0.048 | 0.952 | -0.014 | 0.904 |
| central corpus callosum | pse | TRUE | TRUE | 0.238 | 0.952 | 0.008 | 0.927 |
| left accumbens area | pse | TRUE | TRUE | 0.144 | 0.952 | 0.006 | 0.986 |
| left caudate | pse | TRUE | TRUE | 0.457 | 0.952 | -0.012 | 0.986 |
| left cerebral white matter | pse | TRUE | TRUE | 7.363 | 0.952 | 0.082 | 0.986 |
| right caudate | pse | TRUE | TRUE | 0.333 | 0.952 | -0.001 | 0.986 |
| right inferior lateral ventricle | pse | TRUE | TRUE | 0.108 | 0.952 | -0.001 | 0.986 |
| 4th ventricle | pse | TRUE | TRUE | -0.652 | 0.952 | -0.006 | 0.986 |
| brain stem | pse | TRUE | TRUE | 2.27 | 0.952 | -0.031 | 0.986 |
| left putamen | pse | FALSE | FALSE | 0.417 | 0.952 | -0.003 | 0.986 |
| left ventral diencephalon | pse | FALSE | TRUE | 0.508 | 0.952 | -0.001 | 0.986 |

**Supplementary Table 13:** Regional Fractional Anisotropy Fiber Track Results for PSE & CSE.

| Atlas Track Regions (fa) | Smoke Exposure | Random Slopes | Model Converged | Est. Main effect | Est. Interaction | P-val Main (fdr BY) | P-val Inter (fdr BY) |
| --- | --- | --- | --- | --- | --- | --- | --- |
| left superior corticostriate-frontal cortex only | cse | TRUE | TRUE | 0.000039 | **-0.0000102** | 0.04 | 0 |
| left superior corticostriate-parietal cortex only | cse | TRUE | TRUE | 0.0000568 | **-0.0000138** | 0 | 0 |
| left superior corticostriate | cse | TRUE | TRUE | 0.0000462 | **-0.0000116** | 0.01 | 0 |
| left temporal superior longitudinal fasciculus | cse | TRUE | TRUE | 0.0000823 | **-0.0000103** | 0 | 0 |
| right corticospinal/pyramidal | cse | TRUE | TRUE | 0.0000382 | **-0.000011** | 0.08 | 0 |
| right superior corticostriate-frontal cortex only | cse | TRUE | TRUE | 0.0000425 | **-0.000014** | 0.06 | 0 |
| right superior corticostriate-parietal cortex only | cse | TRUE | TRUE | 0.0000509 | **-0.0000158** | 0.03 | 0 |
| right superior corticostriate | cse | TRUE | TRUE | 0.0000493 | **-0.0000153** | 0.03 | 0 |
| right superior longitudinal fasciculus | cse | TRUE | TRUE | 0.0000474 | **-0.000009** | 0.03 | 0 |
| forceps major | cse | TRUE | TRUE | 0.0000776 | **-0.0000211** | 0 | 0 |
| left cingulate cingulum | cse | FALSE | TRUE | 0.0000714 | **-0.0000128** | 0.04 | 0 |
| left corticospinal/pyramidal | cse | FALSE | TRUE | 0.0000654 | **-0.0000147** | 0 | 0 |
| left fornix | cse | FALSE | TRUE | 0.0000952 | **-0.0000133** | 0 | 0 |
| left fornix, excluding fimbria | cse | FALSE | TRUE | 0.0001114 | **-0.0000161** | 0 | 0 |
| left inferior fronto-occipital fasciculus | cse | FALSE | TRUE | 0.0000804 | **-0.0000114** | 0 | 0 |
| left inferior frontal superior frontal cortex | cse | FALSE | TRUE | 0.0000518 | **-0.0000145** | 0 | 0 |
| left inferior longitudinal fasciculus | cse | FALSE | TRUE | 0.0000624 | **-0.0000108** | 0 | 0 |
| left parietal superior longitudinal fasciculus | cse | FALSE | TRUE | 0.0000602 | **-0.0000066** | 0 | 0 |
| left superior longitudinal fasciculus | cse | FALSE | TRUE | 0.0000778 | **-0.0000084** | 0 | 0 |
| right anterior thalamic radiation | cse | FALSE | TRUE | 0.0000724 | **-0.0000147** | 0 | 0 |
| right inferior fronto-occipital fasciculus | cse | FALSE | TRUE | 0.0000512 | **-0.0000079** | 0.01 | 0 |
| right inferior longitudinal fasciculus | cse | FALSE | TRUE | 0.0000406 | **-0.0000052** | 0.06 | 0 |
| right parietal superior longitudinal fasciculus | cse | FALSE | TRUE | 0.0000445 | **-0.0000091** | 0.03 | 0 |
| corpus callosum | cse | FALSE | TRUE | 0.0000723 | **-0.0000134** | 0 | 0 |
| forceps minor | cse | FALSE | TRUE | 0.0000835 | **-0.0000161** | 0 | 0 |
| right temporal superior longitudinal fasciculus | cse | TRUE | TRUE | 0.0000481 | **-0.0000075** | 0.03 | 0.01 |
| left striatal inferior frontal cortex | cse | TRUE | TRUE | 0.000033 | **-0.0000067** | 0.2 | 0.07 |
| right cingulate cingulum | cse | TRUE | TRUE | 0.0000465 | **-0.0000112** | 0.37 | 0.07 |
| right fornix | cse | TRUE | TRUE | 0.0000525 | **-0.0000077** | 0.01 | 0.08 |
| right striatal inferior frontal cortex | cse | TRUE | TRUE | 0.0000269 | **-0.0000065** | 0.49 | 0.12 |
| right inferior frontal superior frontal cortex | cse | TRUE | TRUE | 0.0000328 | **-0.0000055** | 0.16 | 0.13 |
| right uncinate fasciculus | cse | FALSE | TRUE | 0.0000447 | **-0.0000024** | 0.04 | 0.14 |
| left anterior thalamic radiation | cse | TRUE | TRUE | 0.0000514 | -0.0000031 | 0.03 | 1 |
| left parahpcm cingulum | cse | TRUE | TRUE | 0.0000066 | 0.0000002 | 1 | 1 |
| left uncinate fasciculus | cse | TRUE | TRUE | 0.000072 | -0.0000023 | 0 | 1 |
| right parahpcm cingulum | cse | TRUE | TRUE | 0.0000043 | -0.0000006 | 1 | 1 |
| right fornix, excluding fimbria | cse | TRUE | TRUE | 0.0000372 | -0.0000038 | 0.24 | 1 |
| left striatal inferior frontal cortex | pse | FALSE | TRUE | 0.0000414 | **-0.0000188** | 1 | 0.02 |
| left superior corticostriate-frontal cortex only | pse | TRUE | TRUE | -0.0000365 | 0.0000128 | 1 | 0.15 |
| right superior corticostriate-frontal cortex only | pse | TRUE | TRUE | -0.000072 | 0.0000138 | 1 | 0.15 |
| right inferior longitudinal fasciculus | pse | FALSE | TRUE | -0.0000251 | 0.0000145 | 1 | 0.15 |
| left superior corticostriate | pse | TRUE | TRUE | -0.0000194 | 0.0000105 | 1 | 0.27 |
| right corticospinal/pyramidal | pse | TRUE | TRUE | -0.0000371 | 0.0000114 | 1 | 0.27 |
| left anterior thalamic radiation | pse | FALSE | TRUE | -0.0000747 | 0.0000145 | 1 | 0.27 |
| right uncinate fasciculus | pse | FALSE | TRUE | 0.0000013 | -0.0000141 | 1 | 0.27 |
| forceps minor | pse | FALSE | TRUE | 0.0000317 | -0.0000173 | 1 | 0.27 |
| corpus callosum | pse | FALSE | TRUE | 0.0000214 | -0.0000108 | 1 | 0.33 |
| left parietal superior longitudinal fasciculus | pse | FALSE | TRUE | -0.0000633 | 0.0000112 | 1 | 0.35 |
| right superior corticostriate | pse | FALSE | TRUE | -0.0000505 | 0.0000097 | 1 | 0.35 |
| right anterior thalamic radiation | pse | FALSE | TRUE | -0.0000566 | 0.0000122 | 1 | 0.36 |
| right fornix, excluding fimbria | pse | TRUE | TRUE | -0.0000656 | 0.0000145 | 1 | 0.44 |
| left superior corticostriate-parietal cortex only | pse | TRUE | TRUE | -0.0000121 | 0.0000095 | 1 | 0.49 |
| right parahpcm cingulum | pse | TRUE | TRUE | -0.0000328 | 0.0000145 | 1 | 0.49 |
| left temporal superior longitudinal fasciculus | pse | TRUE | TRUE | -0.0000658 | 0.0000094 | 1 | 0.54 |
| left corticospinal/pyramidal | pse | TRUE | TRUE | -0.0000243 | 0.0000083 | 1 | 0.62 |
| left inferior frontal superior frontal cortex | pse | TRUE | TRUE | -0.00001 | 0.0000087 | 1 | 0.62 |
| right fornix | pse | TRUE | TRUE | -0.0000486 | 0.0000109 | 1 | 0.62 |
| right cingulate cingulum | pse | TRUE | TRUE | -0.0000366 | 0.0000153 | 1 | 0.67 |
| right superior corticostriate-parietal cortex only | pse | TRUE | TRUE | -0.0000348 | 0.0000083 | 1 | 0.76 |
| left parahpcm cingulum | pse | TRUE | TRUE | -0.0000058 | 0.0000066 | 1 | 1 |
| left fornix | pse | TRUE | TRUE | -0.0000086 | 0.0000073 | 1 | 1 |
| left fornix, excluding fimbria | pse | TRUE | TRUE | -0.0000404 | 0.0000056 | 1 | 1 |
| left inferior fronto-occipital fasciculus | pse | TRUE | TRUE | 0.0000016 | -0.0000006 | 1 | 1 |
| left uncinate fasciculus | pse | TRUE | TRUE | -0.000039 | 0.0000022 | 1 | 1 |
| right inferior fronto-occipital fasciculus | pse | TRUE | TRUE | -0.0000147 | 0.0000009 | 1 | 1 |
| right inferior frontal superior frontal cortex | pse | TRUE | TRUE | -0.0000142 | 0.0000038 | 1 | 1 |
| right striatal inferior frontal cortex | pse | TRUE | TRUE | 0.000002 | -0.0000074 | 1 | 1 |
| right superior longitudinal fasciculus | pse | TRUE | TRUE | -0.0000399 | 0.0000057 | 1 | 1 |
| right temporal superior longitudinal fasciculus | pse | TRUE | TRUE | -0.0000373 | 0.0000059 | 1 | 1 |
| forceps major | pse | TRUE | TRUE | 0.0000607 | -0.0000012 | 1 | 1 |
| left cingulate cingulum | pse | FALSE | TRUE | -0.0000421 | -0.0000049 | 1 | 1 |
| left inferior longitudinal fasciculus | pse | FALSE | TRUE | 0.0000129 | -0.0000022 | 1 | 1 |
| left superior longitudinal fasciculus | pse | FALSE | TRUE | -0.000058 | 0.0000047 | 1 | 1 |
| right parietal superior longitudinal fasciculus | pse | FALSE | TRUE | -0.0000296 | -0.0000008 | 1 | 1 |

**Supplementary Table 14:** Regional Mean Diffusivity Fiber Track Results for PSE & CSE.

| Atlas Track Regions (md) | Smoke Exposure | Random Slopes | Model Converged | Est. Main effect | Est. Interaction | P-val Main (fdr BY) | P-val Inter (fdr BY) |
| --- | --- | --- | --- | --- | --- | --- | --- |
| left anterior thalamic radiation | cse | TRUE | TRUE | 0.00001 | **0.0000114** | 1 | 0 |
| left fornix | cse | TRUE | TRUE | 0.0000024 | **0.0000199** | 1 | 0 |
| left fornix, excluding fimbria | cse | TRUE | TRUE | 0.0000109 | **0.0000214** | 1 | 0 |
| left inferior longitudinal fasciculus | cse | TRUE | TRUE | -0.0000175 | **0.000006** | 1 | 0 |
| left parietal superior longitudinal fasciculus | cse | TRUE | TRUE | -0.00001 | **0.0000098** | 1 | 0 |
| left superior corticostriate | cse | TRUE | TRUE | -0.0000107 | **0.0000117** | 1 | 0 |
| left striatal inferior frontal cortex | cse | TRUE | TRUE | 0.0000034 | **0.0000084** | 1 | 0 |
| right anterior thalamic radiation | cse | TRUE | TRUE | 0.0000101 | **0.0000116** | 1 | 0 |
| right cingulate cingulum | cse | TRUE | TRUE | 0.0000059 | **0.0000151** | 1 | 0 |
| right parahpcm cingulum | cse | TRUE | TRUE | 0.0000195 | **0.0000112** | 1 | 0 |
| right fornix, excluding fimbria | cse | TRUE | TRUE | 0.0000254 | **0.0000187** | 1 | 0 |
| right inferior fronto-occipital fasciculus | cse | TRUE | TRUE | -0.0000082 | **0.0000083** | 1 | 0 |
| right inferior longitudinal fasciculus | cse | TRUE | TRUE | -0.0000207 | **0.0000061** | 1 | 0 |
| right parietal superior longitudinal fasciculus | cse | TRUE | TRUE | -0.0000188 | **0.0000117** | 0.98 | 0 |
| right striatal inferior frontal cortex | cse | TRUE | TRUE | -0.0000005 | **0.0000119** | 1 | 0 |
| right uncinate fasciculus | cse | TRUE | TRUE | 0.0000062 | **0.000011** | 1 | 0 |
| corpus callosum | cse | TRUE | TRUE | 0.0000019 | **0.0000125** | 1 | 0 |
| forceps major | cse | TRUE | TRUE | -0.0000119 | **0.0000147** | 1 | 0 |
| left cingulate cingulum | cse | FALSE | TRUE | 0.0000417 | **-0.0000062** | 0.01 | 0 |
| left parahpcm cingulum | cse | FALSE | TRUE | 0.0000531 | **-0.0000071** | 0 | 0 |
| left corticospinal/pyramidal | cse | FALSE | TRUE | 0.0000361 | **-0.0000042** | 0 | 0 |
| left inferior fronto-occipital fasciculus | cse | FALSE | TRUE | 0.0000112 | **-0.0000037** | 1 | 0 |
| left superior corticostriate-parietal cortex only | cse | FALSE | TRUE | 0.0000159 | **-0.0000018** | 1 | 0 |
| left uncinate fasciculus | cse | FALSE | FALSE | 0.0000136 | -0.0000024 | 1 | 0 |
| right corticospinal/pyramidal | cse | FALSE | TRUE | 0.0000376 | **-0.0000051** | 0 | 0 |
| right fornix | cse | FALSE | TRUE | 0.0000688 | **-0.0000106** | 0 | 0 |
| left temporal superior longitudinal fasciculus | cse | FALSE | TRUE | 0.0000025 | **-0.0000014** | 1 | 0.03 |
| forceps minor | cse | FALSE | FALSE | 0.0000324 | -0.0000025 | 0.39 | 0.04 |
| left inferior frontal superior frontal cortex | cse | FALSE | TRUE | -0.0000012 | 0.0000014 | 1 | 0.05 |
| left superior longitudinal fasciculus | cse | FALSE | TRUE | 0.0000022 | -0.0000013 | 1 | 0.05 |
| right superior corticostriate | cse | FALSE | TRUE | 0.0000098 | -0.0000011 | 1 | 0.1 |
| right superior corticostriate-parietal cortex only | cse | FALSE | TRUE | 0.0000093 | -0.0000011 | 1 | 0.11 |
| right superior corticostriate-frontal cortex only | cse | FALSE | TRUE | 0.000006 | -0.0000009 | 1 | 0.28 |
| left superior corticostriate-frontal cortex only | cse | FALSE | TRUE | 0.0000073 | 0.0000005 | 1 | 1 |
| right inferior frontal superior frontal cortex | cse | FALSE | TRUE | 0.0000027 | -0.0000005 | 1 | 1 |
| right superior longitudinal fasciculus | cse | FALSE | TRUE | -0.0000022 | -0.0000002 | 1 | 1 |
| right temporal superior longitudinal fasciculus | cse | FALSE | TRUE | -0.0000053 | -0.0000003 | 1 | 1 |
| left fornix, excluding fimbria | pse | FALSE | FALSE | -0.0000315 | 0.0000163 | 1 | 0.38 |
| forceps major | pse | FALSE | TRUE | -0.0000729 | 0.0000096 | 0.14 | 0.94 |
| left anterior thalamic radiation | pse | TRUE | TRUE | 0.0000155 | 0.0000014 | 1 | 1 |
| left cingulate cingulum | pse | TRUE | TRUE | 0.0000138 | -0.0000047 | 1 | 1 |
| left parahpcm cingulum | pse | TRUE | TRUE | -0.0000292 | 0.0000002 | 1 | 1 |
| left inferior longitudinal fasciculus | pse | TRUE | TRUE | -0.0000049 | -0.0000038 | 1 | 1 |
| left parietal superior longitudinal fasciculus | pse | TRUE | TRUE | -0.0000102 | -0.0000015 | 1 | 1 |
| left temporal superior longitudinal fasciculus | pse | TRUE | TRUE | -0.000011 | -0.0000016 | 1 | 1 |
| right anterior thalamic radiation | pse | TRUE | TRUE | 0.0000008 | -0.0000001 | 1 | 1 |
| right parahpcm cingulum | pse | TRUE | TRUE | -0.0000246 | 0.0000044 | 1 | 1 |
| right superior corticostriate-frontal cortex only | pse | TRUE | TRUE | 0.0000023 | -0.0000019 | 1 | 1 |
| right fornix | pse | TRUE | TRUE | -0.0000577 | 0.0000093 | 0.79 | 1 |
| right fornix, excluding fimbria | pse | TRUE | TRUE | -0.0000259 | 0.000005 | 1 | 1 |
| right inferior fronto-occipital fasciculus | pse | TRUE | TRUE | -0.0000069 | 0.0000039 | 1 | 1 |
| right superior corticostriate-parietal cortex only | pse | TRUE | TRUE | -0.0000364 | 0.0000013 | 1 | 1 |
| right parietal superior longitudinal fasciculus | pse | TRUE | TRUE | -0.0000185 | 0.0000022 | 1 | 1 |
| right superior corticostriate | pse | TRUE | TRUE | -0.000029 | 0.0000014 | 1 | 1 |
| right superior longitudinal fasciculus | pse | TRUE | TRUE | -0.0000185 | 0.0000018 | 1 | 1 |
| right temporal superior longitudinal fasciculus | pse | TRUE | TRUE | -0.0000142 | 0.0000009 | 1 | 1 |
| right uncinate fasciculus | pse | TRUE | TRUE | -0.0000105 | 0.0000069 | 1 | 1 |
| corpus callosum | pse | TRUE | TRUE | -0.0000224 | 0.000003 | 1 | 1 |
| left corticospinal/pyramidal | pse | FALSE | TRUE | -0.0000205 | 0.0000049 | 1 | 1 |
| left superior corticostriate-frontal cortex only | pse | FALSE | TRUE | 0.0000057 | -0.000003 | 1 | 1 |
| left fornix | pse | FALSE | TRUE | -0.0000571 | 0.0000053 | 0.79 | 1 |
| left inferior fronto-occipital fasciculus | pse | FALSE | TRUE | -0.0000181 | 0.0000044 | 1 | 1 |
| left inferior frontal superior frontal cortex | pse | FALSE | TRUE | 0.000011 | -0.0000041 | 1 | 1 |
| left superior corticostriate-parietal cortex only | pse | FALSE | TRUE | -0.0000295 | 0.0000013 | 1 | 1 |
| left superior corticostriate | pse | FALSE | FALSE | -0.0000146 | -0.0000001 | 1 | 1 |
| left striatal inferior frontal cortex | pse | FALSE | TRUE | -0.0000022 | -0.0000018 | 1 | 1 |
| left superior longitudinal fasciculus | pse | FALSE | TRUE | -0.0000187 | -0.0000006 | 1 | 1 |
| left uncinate fasciculus | pse | FALSE | TRUE | 0.0000035 | -0.0000019 | 1 | 1 |
| right cingulate cingulum | pse | FALSE | TRUE | -0.0000279 | -0.0000031 | 1 | 1 |
| right corticospinal/pyramidal | pse | FALSE | TRUE | -0.0000174 | 0.000005 | 1 | 1 |
| right inferior frontal superior frontal cortex | pse | FALSE | TRUE | -0.0000017 | 0.0000026 | 1 | 1 |
| right inferior longitudinal fasciculus | pse | FALSE | TRUE | -0.0000132 | 0.0000036 | 1 | 1 |
| right striatal inferior frontal cortex | pse | FALSE | TRUE | -0.0000159 | 0.0000035 | 1 | 1 |
| forceps minor | pse | FALSE | TRUE | -0.0000317 | -0.0000034 | 1 | 1 |

## Footnotes

1 This robustness test was added post hoc following separate conversa6ons with Anders Fjell and Anders Dale. In our design, principal components increase precision (by reducing variance) yet they do not change our results as our design u6lizes within-site temporal variance; if there were systema6c across-site reasons for recrui6ng individuals from one gene6c ancestry earlier (or later) this control would be warranted. In addi6on, the within-person longitudinal design, also probably helps guard (par6ally) against this. We thank them for their input.

